# Cardiovascular-Kidney-Metabolic Health in US Adults Under the 2026 Multisociety Guideline: Stage Redistribution From 1999 to 2023 and Population Burden Through 2050

**DOI:** 10.64898/2026.06.08.26355220

**Authors:** Fengzhou Fu, Sini Fang, Huanting Liu, Nan Guo, Shicheng Li, Liqiu Yan

## Abstract

**Background:** The 2026 multisociety guideline frames cardiovascular, kidney, and metabolic health as a staged pathway. National surveillance should distinguish early-risk expansion from advanced-disease accumulation because they require different responses.

**Methods:** We analysed 66,553 adults aged 20 years or older across 11 non-overlapping National Health and Nutrition Examination Survey periods from 1999-2000 to August 2021-August 2023. Staging incorporated adiposity, glycaemia, metabolic risk, KDIGO kidney risk, PREVENT 10-year cardiovascular disease risk, and clinical cardiovascular disease. We estimated age-standardised prevalence and secular trends and projected population burden through 2050. Robustness analyses included complete-case estimation, alternative staging thresholds, exclusion of the latest survey period, and rolling temporal validation.

**Findings:** In 2021-2023, 88.5% (95% CI 87.2-89.9) of US adults were in stage 1 or higher, and 62.1% (59.9-64.4) were in stages 2-4. From 1999-2000 to 2021-2023, stage 0 decreased by 4.3 percentage points and stage 1 increased by 7.9 points, whereas stages 3-4 remained stable. Adiposity and diabetes increased, while hypertension and hypertriglyceridaemia declined. The prevalence of stages 2-4 ranged from 54.4% among college graduates to 69.6% among adults with less than a high-school education. Under population ageing alone, 189.5 million adults were projected to be in stages 2-4 by 2050. Rolling temporal validation yielded a mean absolute error of 1.6 percentage points.

**Interpretation:** The distribution of CKM stages in the US population shifted towards earlier metabolic risk without a parallel increase in advanced-stage disease. Policy should combine upstream prevention, equitable access to detection and treatment, and preparation for a growing older population requiring integrated care.

**Funding:** This work was supported by the Special Project for Clinical and Basic Science and Technology Innovation of Guangdong Medical University (GDMULCJC2024112 and GDMULCJC2025138).

**Research in context:** *Evidence before this study:* We searched PubMed from database inception to July 25, 2026, without language restrictions, using combinations of “cardiovascular-kidney-metabolic”, “CKM stage”, “NHANES”, “United States”, “prevalence”, “trend”, “projection”, and “2026 guideline”. Previous studies established the high prevalence of CKM syndrome, quantified overlap among its component conditions, and described demographic, social, and mortality gradients. We identified no national analysis that applied the 2026 multisociety staging framework across all non-overlapping public NHANES periods through August 2023 while distinguishing changes in stage distribution from the demographic translation of prevalence into future absolute care demand.

*Added value of this study:* Across 66,553 adults and 11 survey periods, the increase in any CKM was explained chiefly by a smaller proportion of adults in stage 0 and a larger proportion in stage 1; stages 3-4 remained stable. The analysis also distinguishes relative prevalence from absolute service demand, documents marked contemporary education and income gradients in stages 2-4, and shows that the central interpretation persists in complete-case, alternative-threshold, and latest-cycle-exclusion analyses. Rolling temporal validation produced a mean absolute prediction error of 1.6 percentage points.

*Implications of all the available evidence:* The evidence supports a dual and explicitly equitable response. Upstream policy should reduce entry into the CKM continuum through obesity and diabetes prevention, while health systems expand primary-care, cardiovascular, kidney, and metabolic capacity for an ageing population already meeting stage 2-4 criteria. Stable age-standardised advanced-stage prevalence should not be interpreted as stable workload, particularly when lower educational attainment and income are associated with substantially higher burden.

## Introduction

Cardiovascular-kidney-metabolic (CKM) syndrome describes the biological and clinical continuum from excess or dysfunctional adiposity through metabolic disturbance and kidney injury to high cardiovascular risk and established cardiovascular disease. The 2026 AHA/ACC/ADA/ASN guideline is the first comprehensive clinical practice guideline to organise prevention, detection, and treatment along this continuum.^1^ It builds on the American Heart Association’s original staging advisory.^2^ A companion evidence synopsis set out the mechanistic and therapeutic rationale for treating the three organ systems as an interconnected whole.^3^ Quantitative risk estimation is integral to this approach. The PREVENT framework was developed specifically to estimate total cardiovascular disease risk in the CKM setting.^4^ Its sex-specific, race-free equations incorporate kidney function and estimate both atherosclerotic events and heart failure.^5^ External validation in a nationally representative US sample showed strong discrimination and underscored the importance of evaluating calibration in the population in which the equations are applied.^6^

The high prevalence of CKM syndrome in the USA is already established. A JAMA analysis of NHANES 2011-2020 found that almost nine in ten adults met criteria for stage 1 or higher.^7^ An independent US estimate reached a similar conclusion.^8^ Earlier work documented extensive overlap among cardiac, kidney, and metabolic conditions.^9^ Subsequent studies moved beyond prevalence alone. One national analysis linked advanced stages to socioeconomic disadvantage and higher mortality.^10^ Thirty-year data demonstrated important sex differences in stage distribution and risk.^11^ Social determinants were strongly patterned across stages.^12^ Environmental and social disadvantage also tracked with CKM-related mortality.^13^ Together, these studies establish the scale and inequality of CKM syndrome but do not show where along the staged continuum secular change has occurred.

That distinction matters clinically. Later CKM stages are associated with progressively higher mortality in large population cohorts.^14^ A favourable lifestyle is associated with lower mortality across stages, although substantial residual risk remains in advanced disease.^15^ National data from China show that the staging framework also captures marked gradients in cardiovascular and all-cause mortality outside the USA.^16^ In older adults from the ARIC study, higher CKM stages were accompanied by adverse cardiac remodelling and greater heart-failure risk.^17^ An increase confined to stage 1 would therefore call primarily for earlier prevention, whereas growth in stages 3-4 would signal an expanding population already requiring intensive risk assessment, kidney surveillance, and disease-modifying treatment. US risk-factor trends make either pattern plausible.

Cardiovascular risk factors have changed unevenly across racial, ethnic, and socioeconomic groups.^18^ The proportion of adults with optimal cardiometabolic health has remained low.^19^ The latest national estimates confirm a persistently high prevalence of obesity and severe obesity.^20^

Among adults with diabetes, obesity has increased despite improvements in selected risk-factor targets.^21^ Blood-pressure control has also varied substantially over time.^22^ These opposing trends can be obscured by an overall CKM prevalence near 90%. A second source of uncertainty is demographic: even stable or declining age-standardised prevalence can translate into more patients as the population grows and ages.

We therefore addressed three surveillance questions. First, how did the population distribution across CKM stages change among US adults from 1999-2000 through August 2021-August 2023 under a survey-operationalised implementation of the 2026 guideline? Second, how did the major metabolic, kidney, and cardiovascular components embedded in staging change over time? Third, how might population ageing and alternative risk trajectories translate current stage 2-4 prevalence into future numbers requiring CKM-oriented care? Exploratory analyses examined contemporary socioeconomic gradients, robustness to alternative staging assumptions, and the temporal transportability of the trend model.

## Methods

### Study design and population

We conducted a serial cross-sectional analysis of publicly available NHANES data. NHANES uses a complex, multistage probability design to represent the non-institutionalised civilian US population and combines household interviews, mobile examination centre assessments, laboratory testing, and medication questionnaires. We used 11 non-overlapping survey periods: nine conventional two-year cycles from 1999-2000 through 2015-2016, the combined 2017-March 2020 pre-pandemic release, and August 2021-August 2023. The incomplete 2019-March 2020 sample was not analysed separately because it is not nationally representative; NCHS combined it with 2017-2018 and supplied dedicated survey weights.^23^ The 2021-2023 release was kept separate because field operations resumed with a new sample design and modified procedures.^24^ Weight selection and variance estimation followed the NCHS analytic framework.^25^

Adults aged 20 years or older who participated in the mobile examination component were eligible. The analytic sample comprised 66,553 adults: 4,880 in 1999-2000, 5,411 in 2001-2002, 5,041 in 2003-2004, 4,979 in 2005-2006, 5,935 in 2007-2008, 6,218 in 2009-2010, 5,560 in 2011-2012, 5,769 in 2013-2014, 5,719 in 2015-2016, 9,232 in 2017-March 2020, and 7,809 in 2021-2023. The primary staging analysis did not impose complete-case eligibility for every lower-stage measure. Instead, each participant was assigned the highest stage supported by available observations; analyses of individual components used the measurements and weights required for that component. Cycle-specific sample sizes and stage composition are reported in Supplementary Table 2.

The estimand was the prevalence, in each survey period, of the highest observable CKM stage. An observed higher-stage condition took precedence over missing information for lower stages. We did not impute features that were structurally unavailable in some periods, because imputation cannot recover a guideline component that was never collected. Component-specific denominators are therefore reported, and endpoint sensitivity analyses use laboratory-specific weights. This strategy retains adults with unequivocal clinical cardiovascular disease or very-high kidney risk while making the limits of stage 0 explicit.

All NHANES protocols were approved by the National Center for Health Statistics Research Ethics Review Board, and participants provided written informed consent. The present study used only publicly available, deidentified data and required no additional institutional review board approval or participant consent.

### CKM stage assignment

We assigned the highest observable CKM stage hierarchically. Stage 4 was established clinical cardiovascular disease, defined by self-reported physician diagnosis of coronary heart disease, angina, myocardial infarction, heart failure, or stroke. Atrial fibrillation and peripheral artery disease could not be identified consistently across the 25-year series and were not included.

Among adults without stage 4 disease, stage 3 required either very-high-risk chronic kidney disease on the KDIGO eGFR-UACR matrix^26^ or a PREVENT-estimated 10-year total cardiovascular disease risk of at least 20%. PREVENT was calculated for adults aged 30-79 years without clinical cardiovascular disease using the sex-specific, race-free base equations. Predictors were age, total and HDL cholesterol, systolic blood pressure, diabetes, current smoking, body-mass index, eGFR, antihypertensive treatment, and statin use. Values outside the equation-development ranges were set to the nearest permitted boundary: total cholesterol 130-320 mg/dL, HDL cholesterol 20-100 mg/dL, systolic blood pressure 90-180 mm Hg, eGFR 15-140 mL/min per 1.73m², and body-mass index 18.5-39.9 kg/m². Adults outside the PREVENT age range could still enter stage 3 through very-high kidney risk.

Stage 2 comprised hypertension, diabetes, fasting triglycerides of at least 135 mg/dL, metabolic syndrome, or moderate-to-high KDIGO kidney risk, in the absence of stage 3 or 4 criteria. Metabolic syndrome required at least three of elevated waist circumference, low HDL cholesterol, triglycerides of at least 150 mg/dL, elevated blood pressure or antihypertensive treatment, and prediabetes. Stage 1 comprised excess or dysfunctional adiposity or prediabetes without higher-stage criteria. Adiposity was defined by body-mass index of at least 23 kg/m² for non-Hispanic Asian adults and at least 25 kg/m² for other adults, or the corresponding sex– and ethnicity-specific waist thresholds. Prediabetes was HbA1c 5.7% to less than 6.5% or fasting glucose 100 to less than 126 mg/dL without diabetes. Stage 0 indicated no observed criterion for stages 1-4. eGFR was calculated with the 2021 race-free CKD-EPI creatinine equation.^27^ The joint use of eGFR and UACR reflects their independent associations with cardiovascular and mortality outcomes.^28^ These measures also carry complementary clinical information across acute and chronic kidney disease.^29^ Operational definitions are summarised in Supplementary Table 1.

### Survey estimation and trend analysis

All estimates incorporated NHANES weights, strata, and primary sampling units. For each analysis, we selected the survey weight corresponding to the most restrictive required component, including the special 1999-2002 four-year weights, the 2017-March 2020 pre-pandemic weights, and the 2021-2023 phlebotomy weights for analyses involving blood analytes. Variances and 95% CIs were estimated by Taylor-series linearisation. Prevalence was directly standardised to the 2010 US adult population in age groups 20-39, 40-59, and 60 years or older.

We calculated absolute differences between 1999-2000 and 2021-2023. Because the samples were independent, the variance of each difference was the sum of the two cycle-specific design variances. Long-term trends were estimated with survey-weighted logistic regression; calendar time was scaled per decade, with adjustment for continuous age, sex, and race and ethnicity. We repeated these models for individual stages, CKM components, and stage 2-4 prevalence within sex, age, and race and ethnicity subgroups. One-knot models were used only to screen for departures from a log-linear trend and were not interpreted as formal joinpoint analyses. (Supplementary Tables 6A-6B).

### Population projection scenarios

We used the main-series 2023 US Census national population projections, which apply a cohort-component method to a 2022 population base.^30^ Projections focused on stages 2-4 because these stages identify established metabolic risk, clinically relevant kidney risk, high predicted cardiovascular risk, or clinical cardiovascular disease and therefore define a practical health-service planning population.

For the population-ageing-only scenario, we estimated recent stage 2-4 prevalence within age groups 20-44, 45-64, and 65 years or older and held those rates constant while applying the projected age composition and adult population totals for 2040 and 2050. Projected burden in year y was the sum, across age groups, of projected population multiplied by 2021-2023 prevalence. The trend-continuation scenario extrapolated the adjusted historical log-odds coefficient to 2040 and 2050 before applying the Census population totals. The risk-improvement scenario reduced the age-specific population-ageing prevalence by 10% in 2040 and 20% in 2050. These percentage reductions were planning assumptions defined for scenario analysis, not estimates of treatment efficacy. (Supplementary Table 7).

The 2023 projection baseline differs slightly from the directly age-standardised 2021-2023 estimate because the two quantities answer different questions. The trend analysis standardised prevalence to the 2010 adult age distribution to support comparison across survey periods. The projection baseline instead reweighted contemporary age-specific prevalence to the population structure and total adult population in the 2023 Census series. We therefore treated the 62.1% age-standardised estimate and the 63.7% projection baseline as methodologically distinct, not discordant.

Projection uncertainty was propagated through 20,000 Monte Carlo draws. We sampled the logit-transformed vector of recent age-specific prevalences from its design-based covariance matrix. For trend continuation, each draw also included a normally distributed value of the estimated time coefficient. Age-specific probabilities were then combined with projected Census populations, and the 2.5th and 97.5th percentiles of the simulated distributions defined uncertainty intervals. Historical calibration compared fitted and observed cycle-level stage 2-4 prevalence. (Supplementary Table 8; Supplementary Figure 2).

### Sensitivity, equity, temporal validation, and role of funding source

We assessed robustness in four complementary ways. First, we repeated endpoint and trend analyses in a complete-case subset with body-mass index, waist circumference, HbA1c, fasting glucose, blood pressure, triglycerides, HDL cholesterol, eGFR, UACR, and clinical cardiovascular disease observed; adults eligible for PREVENT additionally required complete risk-equation inputs. These analyses used laboratory-specific weights. Second, we varied the PREVENT threshold defining stage 3 to 15% and 25%. Third, we applied uniform adiposity thresholds—body-mass index of at least 25 kg/m² and waist circumference of at least 88 cm in women or 102 cm in men—regardless of race and ethnicity, recalculated metabolic syndrome, and repeated staging. Fourth, we refitted trend models after omitting August 2021-August 2023. To characterise selection into the complete-case analysis, we compared demographic and clinical characteristics and survey-period composition between the full and complete-case cohorts using standardised differences.

For policy-relevant inequality analyses, we estimated 2021-2023 age-standardised stages 2-4 prevalence across educational attainment, family income-to-poverty ratio, and insurance status. Separate survey-weighted logistic models estimated associations with stage 2-4 after adjustment for age, sex, and race and ethnicity; these analyses were exploratory and were not interpreted causally. (Supplementary Table 12).

We evaluated temporal transportability with rolling-origin validation. Beginning with a model fitted through 2007-2008, each adjusted stages 2-4 trend model was used to predict the next NHANES period, after which the training window was expanded by one period. We summarised mean absolute error, root mean squared error, and maximum absolute error in age-standardised prevalence. A separate post-2016 holdout analysis fitted the model through 2015-2016 and predicted the final two survey periods. These procedures assess short-horizon transportability and do not validate long-range projections to 2050. (Supplementary Table 13).

We repeated endpoint prevalence estimation with laboratory-specific weights and after excluding participants whose PREVENT predictors required boundary truncation. Current-smoking information for 1999-2004 was restored from independently audited NHANES-derived files before PREVENT recalculation; 39 of 8,107 otherwise eligible adults remained without a risk estimate. Analyses used Python 3 with NumPy, pandas, SciPy, and statsmodels. Tests were two-sided. Statistical significance was defined as p<0.05 for the primary stage outcomes, whereas component and subgroup findings were interpreted from effect size, interval width, and consistency. Reporting followed the STROBE statement.^31^ The funder had no role in study design, data acquisition, analysis, interpretation, writing, or the decision to submit. (Supplementary Table 9; Supplementary Figure 3).

## Results

### Population characteristics and contemporary CKM distribution

The 66,553 participants represented 11 non-overlapping NHANES periods. In the pooled sample, 9,240 participants were assigned to stage 0, 13,617 to stage 1, 32,698 to stage 2, 3,193 to stage 3, and 7,805 to stage 4. Higher stages were characterised by older age, higher blood pressure and glycaemia, lower eGFR, and greater albuminuria (table 1). Among PREVENT-eligible adults, the mean 10-year total cardiovascular disease risk was 25.0% in stage 3.

**Table 1:** Selected characteristics of US adults by 2026 guideline-aligned CKM stage, NHANES 1999-2023.

| Characteristic | Overall | Stage 0 | Stage 1 | Stage 2 | Stage 3 | Stage 4 |
| --- | --- | --- | --- | --- | --- | --- |
| Unweighted n | 66,553 | 9,240 | 13,617 | 32,698 | 3,193 | 7,805 |
| Age, years | 47.2 ± 17.1 | 35.1 ± 12.6 | 39.2 ± 13.5 | 49.4 ± 15.6 | 71.2 ± 8.8 | 65.0 ± 13.5 |
| Body mass index, kg/m<sup>2</sup> | 28.81 ± 6.80 | 21.87 ± 2.01 | 29.16 ± 5.38 | 30.18 ± 7.07 | 29.89 ± 6.35 | 30.30 ± 7.13 |
| Waist circumference, cm | 98.4 ± 16.3 | 79.7 ± 6.8 | 97.8 ± 12.5 | 102.1 ± 16.2 | 106.1 ± 15.0 | 105.1 ± 15.9 |
| Systolic blood pressure, mmHg | 122.4 ± 17.9 | 109.4 ± 9.3 | 112.4 ± 9.0 | 127.8 ± 17.2 | 141.9 ± 23.3 | 130.5 ± 21.6 |
| HbA1c, % | 5.58 ± 0.94 | 5.13 ± 0.28 | 5.30 ± 0.35 | 5.69 ± 1.05 | 6.38 ± 1.36 | 6.08 ± 1.22 |
| eGFR, mL/min/1.73 m<sup>2</sup> | 96.9 ± 21.4 | 107.5 ± 16.8 | 104.1 ± 17.1 | 96.2 ± 19.3 | 67.2 ± 23.6 | 77.4 ± 23.8 |
| UACR, mg/g | 34.0 ± 371.0 | 7.3 ± 5.1 | 6.6 ± 4.6 | 29.2 ± 396.1 | 227.2 ± 988.3 | 103.7 ± 517.2 |
| PREVENT 10-year total CVD risk, % | 5.49 ± 6.46 | 1.67 ± 2.27 | 2.31 ± 2.67 | 5.81 ± 4.93 | 25.04 ± 5.58 | — |
| Men | 31,738<br>(48.1%) | 3,995<br>(40.2%) | 5,945<br>(46.4%) | 15,523<br>(49.4%) | 1,953<br>(57.5%) | 4,322<br>(53.2%) |
| Women | 34,815<br>(51.9%) | 5,245<br>(59.8%) | 7,672<br>(53.6%) | 17,175<br>(50.6%) | 1,240<br>(42.5%) | 3,483<br>(46.8%) |
| Adiposity criterion | 45,441<br>(71.3%) | 0 (0.0%) | 12,210<br>(90.5%) | 25,111<br>(79.6%) | 2,620<br>(83.5%) | 5,500<br>(78.8%) |
| Hypertension | 36,625<br>(51.4%) | 0 (0.0%) | 0 (0.0%) | 26,528<br>(79.9%) | 3,052<br>(95.7%) | 7,045<br>(89.4%) |
| Diabetes | 10,713<br>(12.0%) | 0 (0.0%) | 0 (0.0%) | 6,034<br>(14.4%) | 1,756<br>(51.4%) | 2,923<br>(34.1%) |
| Moderate-to-very-high KDIGO kidney risk | 10,732<br>(13.3%) | 0 (0.0%) | 0 (0.0%) | 5,949<br>(16.2%) | 1,772<br>(53.8%) | 3,011<br>(37.2%) |
| Clinical cardiovascular disease | 7,805 (8.9%) | 0 (0.0%) | 0 (0.0%) | 0 (0.0%) | 0 (0.0%) | 7,805<br>(100.0%) |

Data are survey-weighted mean ± SD or unweighted n (survey-weighted %), unless otherwise indicated. UACR was markedly right-skewed; values are presented descriptively and were not used for between-stage inference. PREVENT risk was summarised among eligible adults without clinical cardiovascular disease and is not shown for stage 4. CKM=cardiovascular-kidney-metabolic. CVD=cardiovascular disease. eGFR=estimated glomerular filtration rate. UACR=urine albumin-to-creatinine ratio.

The surveyed population also aged over time. The weighted mean age increased from 46.3 years in 1999-2006 to 48.4 years in 2015-2023, and the proportion aged 60 years or older rose from 22.7% to 29.6%. Across the same broad eras, stage 0 declined from 16.3% to 11.5%, stage 1 increased from 20.9% to 26.0%, and stage 2 remained close to half of the adult population. These descriptive estimates anticipated the cycle-specific age-standardised pattern and motivated separate analyses of prevalence and demographic burden.

In 2021-2023, the age-standardised prevalence of stage 1 or higher was 88.5% (95% CI 87.2-89.9); the prevalence of stages 2-4 was 62.1% (59.9-64.4), and that of stages 3-4 was 11.7% (10.8-12.7). Stage-specific prevalence was 11.5% for stage 0, 26.4% for stage 1, 50.4% for stage 2, 3.0% for stage 3, and 8.7% for stage 4. (Supplementary Table 3; Supplementary Figure 1).

### Redistribution across CKM stages

From 1999-2000 to 2021-2023, any CKM increased by 4.3 percentage points (95% CI 1.8-6.7; p<0.001). Stages 2-4 decreased by 3.6 points (–7.2 to –0.1; p=0.046), whereas stages 3-4 changed little (–0.8 points, –2.7 to 1.1; p=0.411; table 2). The stage-specific results locate the main shift at the entry to the continuum: stage 0 fell by 4.3 points and stage 1 rose by 7.9 points. Stage 2 declined modestly by 2.8 points, while stages 3 and 4 remained stable (figure 1).

**Figure 1:**
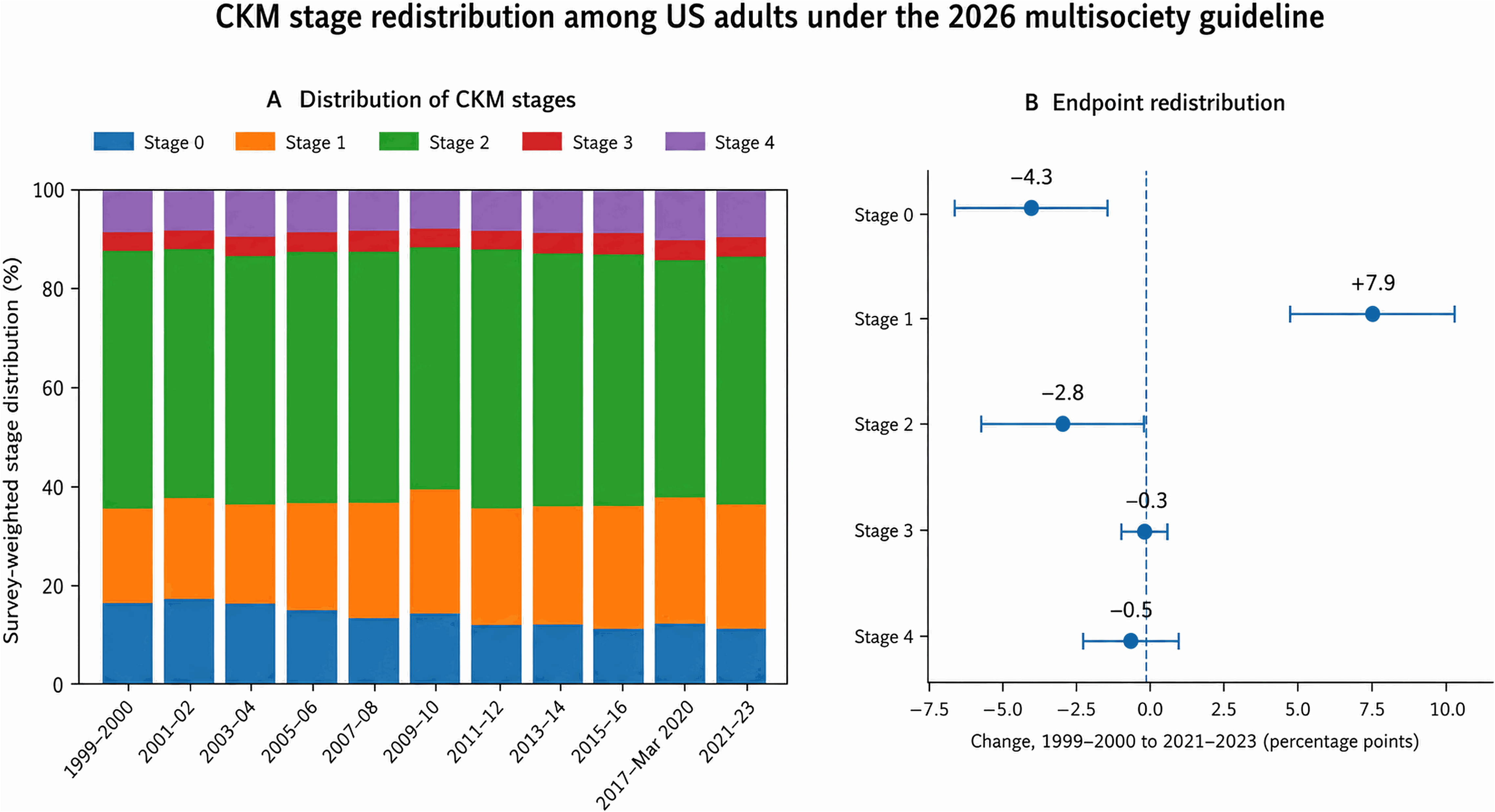
CKM stage redistribution among US adults under the 2026 multisociety guideline. Panel A shows survey-weighted stage composition across 11 non-overlapping NHANES periods. Panel B shows age-standardised changes from 1999-2000 to 2021-2023 with 95% CIs. The dominant change was a decrease in stage 0 and an increase in stage 1, without a significant change in stages 3 or 4.

**Table 2:** Age-standardised CKM prevalence, endpoint redistribution, and adjusted long-term trends.

| Outcome | 1999-2000,<br>% (95% CI) | 2021-2023,<br>% (95% CI) | Absolute change,<br>pp (95% CI) | Adjusted OR per<br>decade (95% CI) | P trend |
| --- | --- | --- | --- | --- | --- |
| Any CKM | 84.3 (82.2-86.3) | 88.5 (87.2-89.9) | +4.3 (1.8-6.7) | 1.23 (1.15-1.31) | <0.001 |
| Stages 2-4 | 65.7 (63.0-68.5) | 62.1 (59.9-64.4) | -3.6 (-7.2 to -0.1) | 0.92 (0.87-0.97) | 0.004 |
| Stages 3-4 | 12.5 (10.9-14.2) | 11.7 (10.8-12.7) | -0.8 (-2.7 to 1.1) | 0.95 (0.89-1.01) | 0.102 |
| Stage 0 | 15.7 (13.7-17.8) | 11.5 (10.1-12.8) | -4.3 (-6.7 to -1.8) | 0.82 (0.76-0.87) | <0.001 |
| Stage 1 | 18.5 (16.3-20.7) | 26.4 (24.8-28.0) | +7.9 (5.2-10.6) | 1.23 (1.17-1.28) | <0.001 |
| Stage 2 | 53.2 (51.4-55.0) | 50.4 (48.3-52.5) | -2.8 (-5.6 to -0.1) | 0.95 (0.92-0.99) | 0.017 |
| Stage 3 | 3.3 (2.8-3.8) | 3.0 (2.6-3.4) | -0.3 (-0.9 to 0.4) | 0.99 (0.92-1.08) | 0.888 |
| Stage 4 | 9.2 (7.8-10.6) | 8.7 (7.8-9.6) | -0.5 (-2.2 to 1.1) | 0.97 (0.90-1.05) | 0.442 |

Age standardisation used the 2010 US adult population. Odds ratios were estimated per decade with adjustment for age, sex, and race and ethnicity and accounted for NHANES strata, primary sampling units, and survey weights. pp=percentage points. OR=odds ratio.

Adjusted models gave the same interpretation. Per decade, the odds of any CKM increased (OR 1.23, 95% CI 1.15-1.31), stage 1 increased (1.23, 1.17-1.28), and stage 0 decreased (0.82, 0.76-0.87). Stages 2-4 declined modestly (0.92, 0.87-0.97), but neither stages 3-4 (0.95, 0.89-1.01) nor stage 3 alone (0.99, 0.92-1.08) showed a significant linear trend. At the population level, the dominant redistribution was a lower proportion in stage 0 and a higher proportion in stage 1, rather than an increasing proportion in stages 3 or 4.

### Divergent component and demographic trends

The components used in staging did not move in parallel (figure 2A). The odds of meeting the adiposity criterion increased by 26% per decade (OR 1.26, 95% CI 1.20-1.33), and diabetes increased by 21% (1.21, 1.14-1.29). Hypertriglyceridaemia declined substantially (0.61, 0.58-0.65), and hypertension declined modestly (0.94, 0.89-0.99). Moderate-to-very-high KDIGO kidney risk (0.98, 0.93-1.03) and clinical cardiovascular disease (0.97, 0.90-1.05) were stable. The opposing component trends explain how stage 1 could expand while stages 2-4 declined modestly. (Supplementary Table 4).

**Figure 2:**
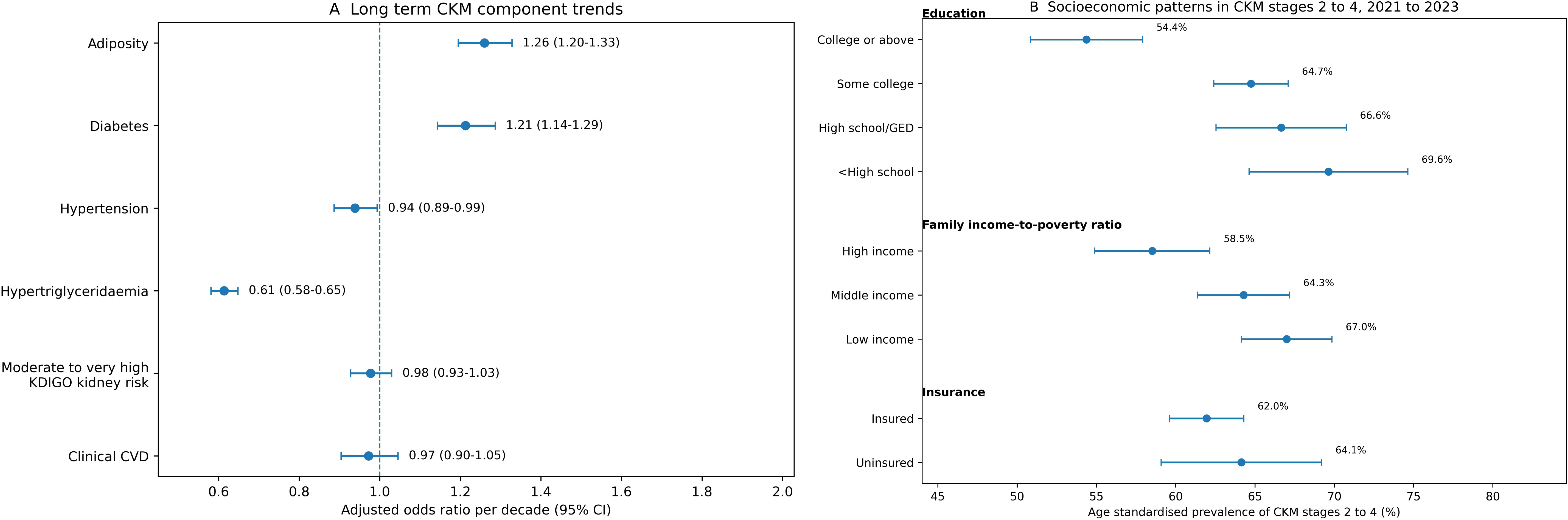
Long-term CKM component trends and contemporary socioeconomic gradients. Panel A shows adjusted odds ratios per decade for major CKM components. Panel B shows age-standardised prevalence of CKM stages 2-4 by educational attainment, family income-to-poverty ratio, and insurance status in August 2021-August 2023. Error bars indicate 95% CIs. Component models were survey-weighted and adjusted for age, sex, and race and ethnicity; socioeconomic prevalence estimates were directly age standardised.

The decline in stages 2-4 was most pronounced among adults aged 60 years or older (OR per decade 0.71, 95% CI 0.63-0.80) and was smaller among those aged 40-59 years (0.90, 0.84-0.98); no linear change was observed at ages 20-39 years. Men and non-Hispanic White adults had modest declines. Estimates in other racial and ethnic groups were compatible with no linear trend but were less precise, particularly where categories were introduced only in later surveys. These subgroup models were descriptive and were not used to infer causal differences (Supplementary Table 5; Supplementary Figure 4).

### Socioeconomic gradients and robustness analyses

In 2021-2023, the prevalence of stages 2-4 showed a clear socioeconomic gradient (figure 2B; Supplementary Table 12). Age-standardised prevalence was 54.4% among adults with a college education or higher, 64.7% among those with some college, 66.6% among high-school graduates, and 69.6% among adults with less than a high-school education. Relative to college graduates, adjusted odds ratios were 1.68 (95% CI 1.33-2.12), 1.85 (1.38-2.48), and 2.17 (1.60-2.94), respectively. Prevalence was 58.5% in the high-income group and 67.0% in the low-income group; the adjusted odds ratio for low versus high income was 1.56 (1.31-1.87). Insurance status was not independently associated with stages 2-4 after demographic adjustment.

The central temporal interpretation was robust, although complete-case estimates were somewhat higher for stages 2-4. Among 25,680 complete-case participants, the adjusted odds of any CKM increased per decade (OR 1.11, 95% CI 1.01-1.23), stages 2-4 declined (0.81, 0.76-0.87), and stages 3-4 remained stable (0.96, 0.88-1.04). The complete-case subset was similar to the full cohort in age, sex, race and ethnicity, body-mass index, diabetes, clinical cardiovascular disease, and survey-period composition (all absolute standardised differences <0.05), but it was modestly enriched for stages 2-4 (71.8% vs 65.7%; standardised difference 0.13; appendix). Lowering or raising the PREVENT threshold changed contemporary stages 3-4 prevalence to 14.4% or 10.2%, respectively, but did not produce a significant linear trend in stage 3. Uniform adiposity thresholds and exclusion of the 2021-2023 period yielded the same qualitative pattern. Rolling-origin temporal validation had a mean absolute error of 1.58 percentage points, a root mean squared error of 1.73 points, and a maximum error of 2.64 points; errors in the two post-2016 holdout periods were 1.38 and 0.89 points. (Supplementary Tables 10-11 and 14).

### Projected population burden through 2050

The projected US adult population increased from 254.2 million in 2023 to 286.2 million in 2050 (12.6%). With contemporary age-specific stage 2-4 prevalence held constant, projected prevalence rose from 63.7% to 66.2% because older adults made up a larger share of the population. The corresponding number of adults with stages 2-4 increased from 162.1 million (95% CI 156.9-167.1) to 189.5 million (184.0-194.9), a rise of 27.5 million or 17.0% (figure 3; table 3).

**Figure 3:**
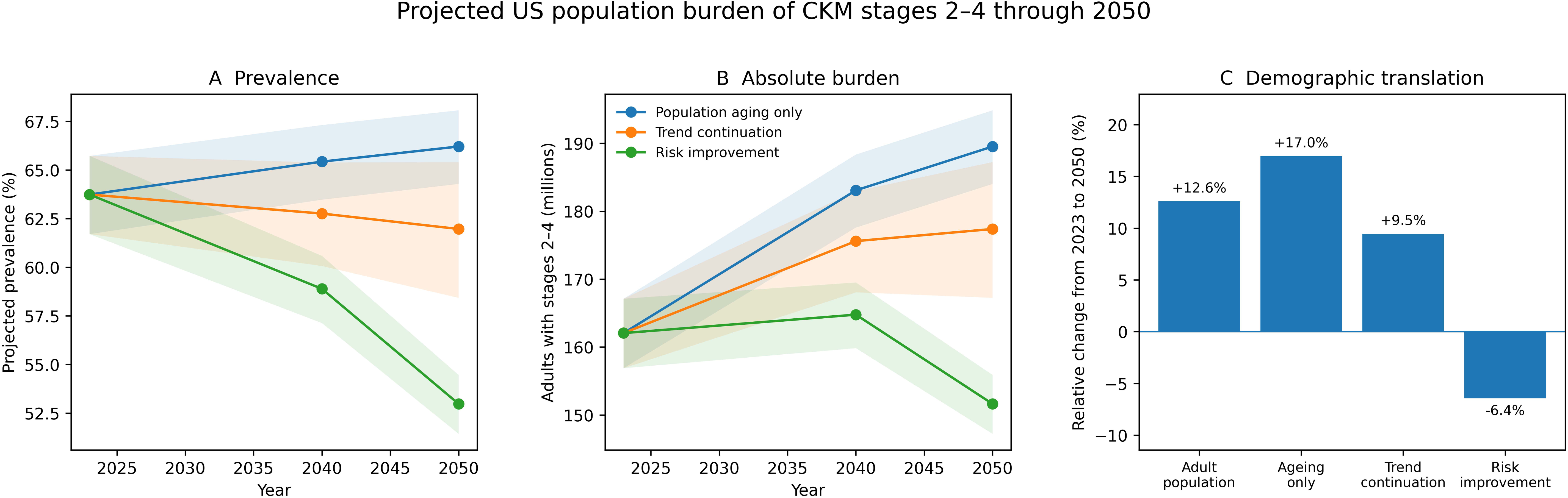
Projected US population burden of CKM stages 2-4 through 2050. Panels A and B show projected prevalence and absolute numbers under population-ageing-only, trend-continuation, and risk-improvement scenarios; shaded regions show 95% uncertainty intervals. Panel C compares relative change in the US adult population and stage 2-4 burden between 2023 and 2050.

**Table 3:** Projected population burden of CKM stages 2-4 through 2050.

| Scenario | Year | Adult population,<br>millions | Stages 2-4 prevalence,<br>% (95% CI) | Adults with stages 2-4,<br>millions (95% CI) | Change from<br>2023, % |
| --- | --- | --- | --- | --- | --- |
| Population ageing only | 2023 | 254.2 | 63.7 (61.7-65.7) | 162.1 (156.9-167.1) | +0.0 |
| Population ageing only | 2040 | 279.8 | 65.4 (63.5-67.3) | 183.1 (177.6-188.3) | +13.0 |
| Population ageing only | 2050 | 286.2 | 66.2 (64.3-68.1) | 189.5 (184.0-194.9) | +17.0 |
| Trend continuation | 2023 | 254.2 | 63.7 (61.7-65.7) | 162.1 (156.9-167.1) | +0.0 |
| Trend continuation | 2040 | 279.8 | 62.8 (60.1-65.4) | 175.6 (168.1-182.9) | +8.4 |
| Trend continuation | 2050 | 286.2 | 62.0 (58.4-65.4) | 177.4 (167.2-187.2) | +9.5 |
| Risk improvement | 2023 | 254.2 | 63.7 (61.7-65.7) | 162.1 (156.9-167.1) | +0.0 |
| Risk improvement | 2040 | 279.8 | 58.9 (57.1-60.6) | 164.8 (159.8-169.5) | +1.7 |
| Risk improvement | 2050 | 286.2 | 53.0 (51.4-54.5) | 151.6 (147.2-155.9) | -6.4 |

Population-ageing-only held recent age-specific prevalence constant. Trend continuation extrapolated the historical adjusted log-odds trend. Risk improvement reduced age-specific prevalence by 10% in 2040 and 20% in 2050 relative to population ageing only. Estimates are scenario exercises rather than forecasts of inevitable disease.

With historical trend continuation, stage 2-4 prevalence declined to 62.0% in 2050, yet the absolute burden still rose to 177.4 million (167.2-187.2), 9.5% above the 2023 estimate. Under risk improvement, prevalence fell to 53.0% and the 2050 burden to 151.6 million (147.2-155.9), 6.4% below the 2023 baseline. Historical fitted prevalence differed from observed cycle estimates by no more than 2.1 percentage points. Complete-case, alternative-definition, and temporal-validation results are reported in the appendix.

## Discussion

In this nationally representative analysis aligned with the observable core of the 2026 multisociety guideline, the main 25-year change was an expansion of early CKM risk rather than a population-wide increase in advanced disease. Stage 0 contracted, stage 1 expanded, and stages 3-4 remained stable. The component analysis helps explain this pattern: adiposity and diabetes became more common, whereas hypertriglyceridaemia and hypertension improved and neither kidney risk nor clinical cardiovascular disease increased. The findings were consistent across complete-case, alternative-threshold, and latest-cycle-exclusion analyses, and short-horizon temporal validation showed modest prediction error. Nevertheless, favourable or stable age-standardised trends at later stages did not translate into lower demand for care. Under population ageing alone, the projected stage 2-4 population increased by about 27 million adults by 2050.

Earlier US studies established the high prevalence of CKM syndrome and the overlap of its component conditions. Our analysis extends that evidence by locating the population-level change predominantly at stage 1 rather than at high predicted risk or clinical cardiovascular disease. That distinction changes the prevention task: a growing population with adiposity or prediabetes calls for earlier intervention, whereas stable stages 3-4 should not be interpreted as evidence that this population will remain at low risk. The serial cross-sectional design cannot determine whether stability at later stages reflects improved treatment, competing survival, or other secular changes.

The divergence among component trends also cautions against judging CKM health from any single conventional risk factor. Albuminuria is a marker of kidney and systemic vascular injury across cardiovascular and metabolic disorders.^32^ It predicts cardiovascular events beyond its role in defining chronic kidney disease.^33^ Yet national testing remains incomplete among adults at risk.^34^

This gap is particularly important in hypertension and diabetes, for which routine UACR measurement remains inconsistent.^35^ Even non-diabetic CKD with lower levels of albuminuria carries appreciable cardiovascular risk.^36^ Recent US data also indicate that albuminuria is common across the CKM spectrum.^37^ Stable kidney-risk prevalence in NHANES therefore cannot be equated with adequate detection or treatment, particularly because a single survey measurement cannot establish chronicity.

The projections expose a second distinction: epidemiological improvement and health-system workload are not synonymous. Under historical trend continuation, the prevalence of stages 2-4 declined, but absolute numbers rose as the population grew and aged. In the ageing-only scenario, the affected population increased by 17%, compared with 12.6% growth in the adult population. Rolling validation supported short-horizon calibration, with a mean absolute error of 1.6 percentage points, but cannot establish accuracy through 2050. Planning should therefore consider the number of people requiring care alongside age-standardised rates.

These findings support two concurrent policy priorities. First, the USA needs population and clinical strategies that reduce entry into the CKM continuum, including healthier food and physical-activity environments, accessible obesity treatment, diabetes-prevention programmes, and systematic assessment of blood pressure, glycaemia, eGFR, and albuminuria. Second, care systems must prepare for older adults already in stages 2-4, including those with pre-heart-failure phenotypes recognised in contemporary heart-failure guidance.^38^

Disease-modifying therapies increasingly cross traditional specialty boundaries. SGLT2 inhibition reduces kidney and cardiovascular events in chronic kidney disease.^39^ Semaglutide reduces major kidney outcomes in type 2 diabetes and chronic kidney disease.^40^ In adults with obesity but without diabetes, semaglutide also reduces cardiovascular events.^41^

Finerenone improves cardiorenal outcomes in diabetic kidney disease.^42^ Its heart-failure benefit is consistent across eGFR and albuminuria strata.^43^ FLOW subgroup data provide additional context for combined GLP-1 receptor agonist and SGLT2 inhibitor use.^44^ Together, these therapies weaken the rationale for fragmented, organ-specific delivery. Their potential to alter future stage distributions will depend on broad, sustained, and equitable uptake; the present serial surveys cannot attribute secular changes to any individual drug class or policy. Prevention should also be judged by whether longer cumulative metabolic exposure in the expanding stage 1 population is translated into later multimorbidity.

National averages also conceal unequal opportunities for prevention and treatment. In 2021-2023, the age-standardised prevalence of stages 2-4 was about 15 percentage points higher among adults with less than a high-school education than among college graduates, and low income remained associated with greater burden after demographic adjustment; insurance status alone did not explain the gradient. These descriptive results identify populations in whom equitable delivery is most needed rather than causal mechanisms. Capacity expansion should therefore combine routine kidney assessment, understandable risk communication, affordable cardiorenal therapies, and community-level obesity and diabetes prevention.

This study has several strengths. It uses 11 non-overlapping, nationally representative periods, including the complete 2017-March 2020 pre-pandemic release and the newest public post-pandemic cycle. It implements PREVENT– and KDIGO-based stage 3 criteria, restores early-cycle smoking inputs before risk estimation, and separates direct age standardisation from demographic projection. Robustness was evaluated with complete-case analyses, alternative PREVENT and adiposity thresholds, exclusion of the newest cycle, and rolling temporal validation. The projection analysis states its Census inputs, age strata, scenario equations, uncertainty procedure, and calibration performance rather than presenting future counts as opaque forecasts.

Several limitations define the interpretation. NHANES is serial cross-sectional, so individual progression and regression cannot be observed. The classification is a survey-operationalised implementation of the guideline’s observable core, not a complete clinical assessment. NT-proBNP is strongly associated with mortality in US adults.^45^ Long-term changes in NT-proBNP track heart-failure and mortality risk.^46^ Its expected range also varies substantially by age and sex.^47^ High-sensitivity troponin carries prognostic information in general populations.^48^ Stored NHANES specimens show that different high-sensitivity troponin assays provide non-identical risk information.^49^ These biomarkers, cardiac imaging, and coronary calcium were not consistently available. Atrial fibrillation and peripheral artery disease—both recognised in contemporary cardiovascular guidance—could not be included across all cycles.^50^ Stage 3 and stage 4 prevalence is therefore likely conservative. Clinical cardiovascular disease was self-reported, and a single eGFR or UACR measurement cannot establish chronic kidney disease persisting for at least 3 months. PREVENT was not estimated outside ages 30-79 years. The projections depend on Census assumptions, extrapolation of a historical log-odds trend, and deliberately simple risk-improvement scenarios; they should be read as planning ranges rather than forecasts of inevitable disease. The complete-case subset was demographically similar to the full cohort but modestly enriched for stages 2-4, so complete-case estimates are best interpreted as a robustness check rather than an alternative national prevalence estimate. Temporal validation covered subsequent NHANES cycles rather than external populations and cannot establish long-range forecast validity.

In conclusion, the distribution of CKM stages in the US population shifted towards early metabolic risk without a uniform increase in advanced-stage disease. This pattern was robust to alternative analytic assumptions, but the contemporary burden was strongly graded by education and income. Demographic ageing is nevertheless likely to enlarge the population requiring stage 2-4 care, even if age-standardised prevalence stabilises or declines. A credible CKM strategy must therefore combine upstream metabolic prevention and equitable access with deliberate expansion of integrated cardiovascular, kidney, metabolic, and primary-care capacity.

## Contributors

FZF and SNF contributed equally. FZF, SNF, SCL, and LQY conceived and designed the study. FZF, SNF, and HTL acquired, harmonised, and analysed the data. FZF, HTL, NG, and LQY accessed and verified the underlying data and independently reproduced the primary numerical results. FZF and SNF drafted the manuscript. NG, SCL, and LQY interpreted the findings and critically revised the manuscript. SCL and LQY supervised the work; LQY obtained funding and had final responsibility for the decision to submit. All authors approved the final manuscript and accept responsibility for the integrity of the work.

## Data sharing

NHANES public-use data are available from the National Center for Health Statistics. Versioned analytic code, derived-variable specifications, aggregate output tables, figure-source data, and a file manifest are included in the accompanying reproducibility archive.

## Acknowledgements

We thank the participants and staff of NHANES. Generative artificial intelligence tools were used only to support code drafting, language editing, and document formatting. The authors independently verified all analytical code, numerical outputs, references, and scientific interpretations; no generative AI system created or altered the source NHANES data.

## References

1. Ndumele CE, Rodriguez F, Dixon DL, et al. 2026 AHA/ACC/ADA/ASN guideline for the prevention, detection, evaluation, and management of cardiovascular-kidney-metabolic syndrome. Circulation 2026; published online June 9. 10.1161/CIR.0000000000001453.

2. Ndumele CE, Rangaswami J, Chow SL, et al. Cardiovascular-kidney-metabolic health: a presidential advisory from the American Heart Association. Circulation 2023; 148: 1606–35.

3. Ndumele CE, Neeland IJ, Tuttle KR, et al. A synopsis of the evidence for the science and clinical management of cardiovascular-kidney-metabolic syndrome. Circulation 2023; 148: 1636–64.

4. Khan SS, Coresh J, Pencina MJ, et al. Novel prediction equations for absolute risk assessment of total cardiovascular disease incorporating cardiovascular-kidney-metabolic health: a scientific statement from the American Heart Association. Circulation 2023; 148: 1982–2004.

5. Khan SS, Matsushita K, Sang Y, et al. Development and validation of the American Heart Association’s PREVENT equations. Circulation 2024; 149: 430–49.

6. Scheuermann B, Brown A, Colburn T, et al. External validation of the American Heart Association PREVENT cardiovascular disease risk equations. JAMA Netw Open 2024; 7: e2438311.

7. Aggarwal R, Ostrominski JW, Vaduganathan M. Prevalence of cardiovascular-kidney-metabolic syndrome stages in US adults, 2011-2020. JAMA 2024; 331: 1858–60.

8. Minhas AMK, Mathew RO, Sperling LS, et al. Prevalence of the cardiovascular-kidney-metabolic syndrome in the United States. J Am Coll Cardiol 2024; 83: 1824–26.

9. Ostrominski JW, Arnold SV, Butler J, et al. Prevalence and overlap of cardiac, renal, and metabolic conditions in US adults, 1999-2020. JAMA Cardiol 2023; 8: 1050–60.

10. Kim JE, Joo J, Kuku KO, et al. Prevalence, disparities, and mortality of cardiovascular-kidney-metabolic syndrome in US adults, 2011-2018. Am J Med 2025; 138: 970–979.e7.

11. Ji H, Sabanayagam C, Matsushita K, et al. Sex differences in cardiovascular-kidney-metabolic syndrome: 30-year US trends and mortality risks. Arterioscler Thromb Vasc Biol 2025; 45: 157–61.

12. Zhu R, Wang R, He J, et al. Prevalence of cardiovascular-kidney-metabolic syndrome stages by social determinants of health. JAMA Netw Open 2024; 7: e2445309.

13. Vieira de Oliveira Salerno PR, Cotton A, Elgudin YE, et al. Social and environmental determinants of health and cardio-kidney-metabolic syndrome-related mortality. JAMA Netw Open 2024; 7: e2435783.

14. Tsai MK, Kao JTW, Wong CS, et al. Cardiovascular-kidney-metabolic syndrome and all-cause and cardiovascular mortality: a retrospective cohort study. PLoS Med 2025; 22: e1004629.

15. Wang M, Qiao Y, Lin R, et al. Association between cardiovascular-kidney-metabolic syndrome, lifestyle, and all-cause and cause-specific mortality: a prospective cohort study. EClinicalMedicine 2025; 90: 103596.

16. Zheng C, Cai A, Sun M, et al. Prevalence and mortality of cardiovascular-kidney-metabolic syndrome in China: a nationwide population-based study. JACC Asia 2025; 5: 898–910.

17. Lassen MCH, Ostrominski JW, Claggett BL, et al. Cardiovascular-kidney-metabolic syndrome stages, echocardiographic characteristics, and heart failure risk: the Atherosclerosis Risk in Communities Study. Circulation 2026; 153: 1268–80.

18. He J, Zhu Z, Bundy JD, Dorans KS, Chen J, Hamm LL. Trends in cardiovascular risk factors in US adults by race and ethnicity and socioeconomic status, 1999-2018. JAMA 2021; 326: 1286–98.

19. O’Hearn M, Lauren BN, Wong JB, Kim DD, Mozaffarian D. Trends and disparities in cardiometabolic health among US adults, 1999-2018. J Am Coll Cardiol 2022; 80: 138–51.

20. Emmerich SD, Fryar CD, Stierman B, Ogden CL. Obesity and severe obesity prevalence in adults: United States, August 2021-August 2023. NCHS Data Brief 2024; 508: 1–8.

21. Hu G, Ding J, Ryan DH. Trends in obesity prevalence and cardiometabolic risk factor control in US adults with diabetes, 1999-2020. Obesity (Silver Spring) 2023; 31: 841–51.

22. Muntner P, Hardy ST, Fine LJ, et al. Trends in blood pressure control among US adults with hypertension, 1999-2000 to 2017-2018. JAMA 2020; 324: 1190–200.

23. National Center for Health Statistics. National Health and Nutrition Examination Survey, 2017-March 2020 pre-pandemic file: sample design, estimation, and analytic guidelines. Hyattsville, MD: NCHS, 2022. https://wwwn.cdc.gov/nchs/nhanes/analyticguidelines.aspx (accessed July 25, 2026).

24. National Center for Health Statistics. National Health and Nutrition Examination Survey analytic guidance and brief overview for the August 2021-August 2023 data files. Hyattsville, MD: NCHS, 2024. https://wwwn.cdc.gov/nchs/nhanes/analyticguidelines.aspx (accessed July 25, 2026).

25. National Center for Health Statistics. NHANES tutorials: weighting module. https://wwwn.cdc.gov/nchs/nhanes/tutorials/weighting.aspx (accessed July 25, 2026).

26. Kidney Disease: Improving Global Outcomes CKD Work Group. KDIGO 2024 clinical practice guideline for the evaluation and management of chronic kidney disease. Kidney Int 2024; 105 (suppl 4S): S117–314.

27. Inker LA, Eneanya ND, Coresh J, et al. New creatinine– and cystatin C-based equations to estimate GFR without race. N Engl J Med 2021; 385: 1737–49.

28. Grams ME, Coresh J, Matsushita K, et al. Estimated glomerular filtration rate, albuminuria, and adverse outcomes: an individual-participant data meta-analysis. JAMA 2023; 330: 1266–77.

29. Levey AS, Grams ME, Inker LA. Uses of GFR and albuminuria level in acute and chronic kidney disease. N Engl J Med 2022; 386: 2120–28.

30. US Census Bureau. 2023 national population projections tables: main series. https://www.census.gov/data/tables/2023/demo/popproj/2023-summary-tables.html (accessed July 25, 2026).

31. von Elm E, Altman DG, Egger M, Pocock SJ, Gøtzsche PC, Vandenbroucke JP. The Strengthening the Reporting of Observational Studies in Epidemiology (STROBE) statement. Lancet 2007; 370: 1453–57.

32. Claudel SE, Verma A. Albuminuria in cardiovascular, kidney, and metabolic disorders: a state-of-the-art review. Circulation 2025; 151: 716–32.

33. Barzilay JI, Farag YMK, Durthaler J. Albuminuria: an underappreciated risk factor for cardiovascular disease. J Am Heart Assoc 2024; 13: e030131.

34. Chu CD, Powe NR, Shlipak MG, et al. Estimated prevalence and testing for albuminuria in US adults at risk for chronic kidney disease. JAMA Netw Open 2023; 6: e2326230.

35. Shin JI, Chang AR, Grams ME, et al. Albuminuria testing in hypertension and diabetes: an individual-participant data meta-analysis in a global consortium. Hypertension 2021; 78: 1042–52.

36. Shulman R, Yang W, Cohen DL, Reese PP, Cohen JB; CRIC Study Investigators. Cardiovascular and kidney outcomes of non-diabetic chronic kidney disease by albuminuria severity: findings from the CRIC Study. Am J Kidney Dis 2024; 84: 742–750.e1.

37. Kibbi R, Huang X, Krishnan V, et al. Albuminuria prevalence among US adults with cardiovascular-kidney-metabolic syndrome. J Am Heart Assoc 2026; 15: e045697.

38. Heidenreich PA, Bozkurt B, Aguilar D, et al. 2022 AHA/ACC/HFSA guideline for the management of heart failure. Circulation 2022; 145: e895–e1032.

39. Herrington WG, Staplin N, Wanner C, et al. Empagliflozin in patients with chronic kidney disease. N Engl J Med 2023; 388: 117–27.

40. Perkovic V, Tuttle KR, Rossing P, et al. Effects of semaglutide on chronic kidney disease in patients with type 2 diabetes. N Engl J Med 2024; 391: 109–21.

41. Lincoff AM, Brown-Frandsen K, Colhoun HM, et al. Semaglutide and cardiovascular outcomes in obesity without diabetes. N Engl J Med 2023; 389: 2221–32.

42. Agarwal R, Filippatos G, Pitt B, et al. Cardiovascular and kidney outcomes with finerenone in patients with type 2 diabetes and chronic kidney disease: the FIDELITY pooled analysis. Eur Heart J 2022; 43: 474–84.

43. Filippatos G, Anker SD, Pitt B, et al. Finerenone and heart failure outcomes by kidney function and albuminuria in chronic kidney disease and diabetes. JACC Heart Fail 2022; 10: 860–70.

44. Mann JFE, Rossing P, Bakris G, et al. Effects of semaglutide with and without concomitant SGLT2 inhibitor use in participants with type 2 diabetes and chronic kidney disease in the FLOW trial. Nat Med 2024; 30: 2849–56.

45. Echouffo-Tcheugui JB, Zhang S, Daya N, et al. NT-proBNP and all-cause and cardiovascular mortality in US adults: a prospective cohort study. J Am Heart Assoc 2023; 12: e029110.

46. Jia X, Al Rifai M, Hoogeveen R, et al. Association of long-term change in N-terminal pro-B-type natriuretic peptide with incident heart failure and death. JAMA Cardiol 2023; 8: 222–30.

47. Welsh P, Campbell RT, Mooney L, et al. Reference ranges for NT-proBNP and risk factors for higher NT-proBNP concentrations in a large general population cohort. Circ Heart Fail 2022; 15: e009427.

48. Aimo A, Georgiopoulos G, Panichella G, et al. High-sensitivity troponins for outcome prediction in the general population: a systematic review and meta-analysis. Eur J Intern Med 2022; 98: 61–68.

49. McEvoy JW, Wang D, Tang O, et al. Four high-sensitivity troponin assays and mortality in US adults with cardiovascular disease: NHANES 1999-2004. Am J Prev Cardiol 2024; 17: 100631.

50. Gornik HL, Aronow HD, Goodney PP, et al. 2024 ACC/AHA guideline for the management of lower extremity peripheral artery disease. Circulation 2024; 149: e1313–e1410.

