## Supplementary Material for "Cardiovascular-Kidney-Metabolic Health in US Adults Under the 2026 Multisociety Guideline: Stage Redistribution From 1999 to 2023 and Population Burden Through 2050"

**Short title:** CKM stage redistribution and population burden

Fengzhou Fu,[a,b,#] Sini Fang,[b,#] Huanting Liu,[a,b] Nan Guo,[c] Shicheng Li,[a,b,*] Liqiu Yan,[a,b,*]

[a]Division of Cardiovascular Epidemiology, Dongguan Cardiovascular Research Institute, Dongguan, 523326, Guangdong, China

[b]Department of Cardiology, The Affiliated Dongguan Songshan Lake Central Hospital, Guangdong Medical University, Dongguan, Guangdong, China

[c]Department of Cardiology, Cangzhou Central Hospital, Hebei Medical University, Cangzhou, Hebei, China

*Corresponding author. Department of Cardiology, Division of Cardiovascular Epidemiology, Dongguan Cardiovascular Research Institute, Dongguan, 523326, Guangdong, China.

### These authors contributed equally.

#### Appendix contents

Supplementary methods

Supplementary Table 1. Survey-operationalised 2026 CKM stage definitions

Supplementary Table 2. Cycle-specific sample sizes and stage composition

Supplementary Table 3. Cycle-specific age-standardised prevalence

Supplementary Table 4. CKM component trend models

Supplementary Table 5. Stage 2–4 subgroup trends

Supplementary Tables 6A–6B. Exploratory one-knot trend screening

Supplementary Table 7. Projection inputs and scenario equations

Supplementary Table 8. Projection model calibration

Supplementary Table 9. Sensitivity analyses

Supplementary Table 10. Complete-case and alternative-definition endpoint estimates

Supplementary Table 11. Sensitivity analyses of long-term trends

Supplementary Table 12. Socioeconomic gradients in CKM stages 2–4, 2021–2023

Supplementary Table 13. Rolling-origin temporal validation

Supplementary Table 14. Comparison of the full analytic cohort and complete-case subset

Supplementary Figures 1–4

#### Supplementary methods

Variable harmonisation and staging. Within each survey period, we mapped variable names, units, laboratory fields, and questionnaire items before deriving the staging criteria. The algorithm assigned the highest stage supported by observed information; a lower-stage component did not need to be present when a higher-stage criterion was documented. This maximum-observed strategy retains adults with unequivocal clinical cardiovascular disease or very-high kidney risk when a lower-stage laboratory measure is missing. Because stage 0 is conditional on the measurements observed, endpoint sensitivity analyses used laboratory-specific weights and alternative PREVENT predictor handling.

Survey weights. Cycle-specific examination weights were used when stage estimation combined examination and questionnaire variables. For August 2021–August 2023 analyses involving blood analytes, phlebotomy weights were applied in sensitivity analysis. The 1999–2002 period used the NCHS four-year weights, and 2017–March 2020 used the dedicated pre-pandemic weights. Design strata and primary sampling units were made unique across pooled periods before variance estimation.

PREVENT recalculation. Early-cycle current-smoking data were restored from independently audited NHANES-derived source files before PREVENT estimation. The official sex-specific base equations were then implemented. Among 8,107 otherwise PREVENT-eligible adults in 1999–2004, smoking status was available for 8,068; 39 remained without a risk estimate. The primary analysis truncated continuous predictors to the equation-development ranges specified in the main manuscript; a sensitivity analysis excluded records requiring truncation.

Projection uncertainty. For age groups 20–44, 45–64, and 65 years or older, we transformed the design-based covariance matrix of 2021–2023 prevalence to the logit scale and generated 20,000 draws from the corresponding multivariate normal distribution. For trend continuation, each simulation also included a normal draw of the survey-weighted time coefficient for the elapsed decades. Age-specific probabilities were combined with Census age shares and adult population totals; percentile intervals were calculated from the simulated distribution.

Complete-case and alternative definitions. Complete-case analyses required body-mass index, waist circumference, HbA1c, fasting glucose, blood pressure, triglycerides, HDL cholesterol, eGFR, UACR, and clinical cardiovascular disease; PREVENT-eligible adults additionally required complete risk-equation inputs. Laboratory-specific survey weights were used. Alternative definitions used PREVENT stage 3 thresholds of 15% and 25%, and a uniform adiposity definition of body-mass index ≥25 kg/m² or waist circumference ≥88 cm in women and ≥102 cm in men regardless of race and ethnicity. Metabolic syndrome and CKM stage were recalculated under the uniform-threshold analysis. Long-term models were also repeated after excluding August 2021–August 2023. To characterise selection, we compared unweighted demographic and clinical characteristics and survey-period composition between the full and complete-case cohorts using standardised differences.

Socioeconomic gradients. In August 2021–August 2023, stages 2–4 prevalence was directly standardised by age within categories of educational attainment, family income-to-poverty ratio, and health insurance. Separate survey-weighted logistic models estimated odds ratios after adjustment for continuous age, sex, and race and ethnicity. These exploratory analyses describe unequal burden but do not identify causal effects of education, income, or insurance.

Temporal validation. Rolling-origin validation began with a model trained through 2007–2008. The adjusted stages 2–4 trend model was used to predict the next survey period, the training window was then expanded, and the process continued through August 2021–August 2023. Predicted probabilities were directly age standardised using the same 2010 adult weights as the observed estimates. We report mean absolute error, root mean squared error, and maximum absolute error. A separate model trained through 2015–2016 predicted the final two periods. These analyses evaluate short-horizon transportability rather than the validity of extrapolation to 2050.

#### Supplementary Table 1: Survey-operationalised 2026 CKM stage definitions

| Stage | Operational definition |
|---|---|
| Stage 0 | No observed stage 1–4 criterion. |
| Stage 1 | Adiposity (Asian BMI ≥23 kg/m² or waist ≥80 cm in women/≥90 cm in men; other BMI ≥25 kg/m² or waist ≥88 cm in women/≥102 cm in men) or prediabetes (HbA1c 5·7% to <6·5% or fasting glucose 100 to <126 mg/dL), without higher-stage criteria. |
| Stage 2 | Hypertension, diabetes, fasting triglycerides ≥135 mg/dL, metabolic syndrome, or moderate/high KDIGO kidney risk, without stage 3 or 4. |
| Stage 3 | No clinical cardiovascular disease and either PREVENT 10-year total CVD risk ≥20% or very-high KDIGO kidney risk. |
| Stage 4 | Self-reported coronary heart disease, angina, myocardial infarction, heart failure, or stroke. |

Cardiac imaging, biomarkers, atrial fibrillation, and peripheral artery disease were unavailable consistently and were not included.

#### Supplementary Table 2: Cycle-specific sample size and survey-weighted CKM stage composition

| Cycle | N | Stage 0, % | Stage 1, % | Stage 2, % | Stage 3, % | Stage 4, % |
|---|---|---|---|---|---|---|
| 1999-2000 | 4880 | 16.7 | 19.5 | 52.2 | 3.1 | 8.6 |
| 2001-2002 | 5411 | 17.2 | 21.2 | 50.2 | 3.2 | 8.3 |
| 2003-2004 | 5041 | 16.4 | 20.6 | 50.2 | 3.1 | 9.7 |
| 2005-2006 | 4979 | 15.1 | 22.4 | 50.5 | 3.3 | 8.8 |
| 2007-2008 | 5935 | 13.3 | 24.0 | 50.6 | 3.6 | 8.6 |
| 2009-2010 | 6218 | 14.2 | 25.5 | 48.9 | 3.4 | 8.0 |
| 2011-2012 | 5560 | 12.0 | 24.6 | 51.6 | 3.1 | 8.6 |
| 2013-2014 | 5769 | 12.2 | 24.8 | 50.5 | 3.7 | 8.7 |
| 2015-2016 | 5719 | 11.2 | 25.9 | 50.3 | 3.9 | 8.6 |
| 2017-March2020 | 9232 | 12.1 | 26.5 | 47.8 | 3.5 | 10.0 |
| 2021-2023 | 7809 | 11.1 | 25.5 | 50.3 | 3.6 | 9.6 |

Stage proportions sum to 100% within rounding.

#### Supplementary Table 3: Cycle-specific age-standardised CKM prevalence, % (95% CI)

| Cycle | Any CKM (stages 1-4) | CKM stages 2-4 | CKM stages 3-4 | Stage 0 | Stage 1 | Stage 2 | Stage 3 | Stage 4 |
|---|---|---|---|---|---|---|---|---|
| 1999-2000 | 84·3 (82·2–86·3) | 65·7 (63·0–68·5) | 12·5 (10·9–14·2) | 15·7 (13·7–17·8) | 18·5 (16·3–20·7) | 53·2 (51·4–55·0) | 3·3 (2·8–3·8) | 9·2 (7·8–10·6) |
| 2001-2002 | 83·5 (82·4–84·5) | 63·1 (61·1–65·2) | 12·8 (11·8–13·8) | 16·5 (15·5–17·6) | 20·4 (18·8–21·9) | 50·3 (48·3–52·3) | 3·7 (3·2–4·2) | 9·1 (8·0–10·2) |
| 2003-2004 | 84·2 (82·9–85·6) | 64·2 (61·8–66·7) | 13·9 (12·5–15·2) | 15·8 (14·4–17·1) | 20·0 (18·3–21·7) | 50·4 (48·6–52·2) | 3·4 (2·9–3·9) | 10·5 (8·9–12·0) |
| 2005-2006 | 85·3 (83·8–86·9) | 63·5 (61·5–65·4) | 13·0 (11·9–14·0) | 14·7 (13·1–16·2) | 21·9 (19·9–23·8) | 50·5 (48·4–52·7) | 3·6 (3·0–4·3) | 9·3 (8·5–10·2) |
| 2007-2008 | 87·0 (86·0–87·9) | 63·4 (61·9–64·9) | 12·8 (11·5–14·1) | 13·0 (12·1–14·0) | 23·6 (22·0–25·2) | 50·6 (49·1–52·2) | 3·8 (3·6–4·1) | 8·9 (7·7–10·2) |
| 2009-2010 | 85·9 (84·6–87·2) | 60·6 (59·0–62·1) | 11·6 (10·8–12·4) | 14·1 (12·8–15·4) | 25·3 (23·1–27·6) | 48·9 (47·5–50·4) | 3·5 (3·0–3·9) | 8·2 (7·2–9·1) |
| 2011-2012 | 87·9 (86·1–89·7) | 63·2 (60·9–65·4) | 11·6 (10·5–12·8) | 12·1 (10·3–13·9) | 24·7 (22·9–26·5) | 51·5 (49·5–53·5) | 3·1 (2·3–3·9) | 8·5 (7·8–9·3) |
| 2013-2014 | 87·7 (86·3–89·1) | 62·6 (60·5–64·7) | 12·0 (11·0–13·1) | 12·3 (10·9–13·7) | 25·1 (23·5–26·7) | 50·5 (48·5–52·5) | 3·6 (3·1–4·1) | 8·5 (7·5–9·5) |
| 2015-2016 | 88·6 (86·9–90·2) | 62·2 (59·8–64·6) | 11·8 (10·7–12·9) | 11·4 (9·8–13·1) | 26·3 (24·4–28·2) | 50·4 (48·6–52·2) | 3·6 (2·9–4·3) | 8·2 (7·5–8·9) |
| 2017-March2020 | 87·6 (86·2–89·0) | 60·4 (57·9–62·8) | 12·4 (11·4–13·3) | 12·4 (11·0–13·8) | 27·2 (25·7–28·8) | 48·0 (45·5–50·4) | 3·1 (2·6–3·5) | 9·3 (8·4–10·2) |
| 2021-2023 | 88·5 (87·2–89·9) | 62·1 (59·9–64·4) | 11·7 (10·8–12·7) | 11·5 (10·1–12·8) | 26·4 (24·8–28·0) | 50·4 (48·3–52·5) | 3·0 (2·6–3·4) | 8·7 (7·8–9·6) |

Direct standardisation used the 2010 US adult age distribution.

#### Supplementary Table 4: Adjusted long-term trends in CKM components

| Outcome | OR (95% CI) | P | Unweighted_n | Events |
|---|---|---|---|---|
| Adiposity criterion | 1·26 (1·20–1·33) | <0·001 | 66553 | 45441 |
| Hypertension | 0·94 (0·89–0·99) | 0·029 | 66553 | 36625 |
| Diabetes | 1·21 (1·14–1·29) | <0·001 | 66553 | 10713 |
| Hypertriglyceridemia | 0·61 (0·58–0·65) | <0·001 | 66553 | 8578 |
| Moderate-to-very-high KDIGO kidney risk | 0·98 (0·93–1·03) | 0·384 | 66553 | 10732 |
| Clinical cardiovascular disease | 0·97 (0·90–1·05) | 0·442 | 66553 | 7805 |

Odds ratios are per decade and adjusted for age, sex, and race and ethnicity.

#### Supplementary Table 5: Adjusted trends in CKM stages 2–4 by subgroup

| Subgroup_variable | Subgroup | OR (95% CI) | P | N | Events |
|---|---|---|---|---|---|
| Sex | Men | 0·89 (0·83–0·96) | 0·002 | 31738 | 21798 |
| Sex | Women | 0·95 (0·89–1·01) | 0·092 | 34815 | 21898 |
| Age group | 20-39 | 1·00 (0·93–1·08) | 0·957 | 22025 | 8463 |
| Age group | 40-59 | 0·90 (0·84–0·98) | 0·010 | 20665 | 13986 |
| Age group | 60+ | 0·71 (0·63–0·80) | <0·001 | 23863 | 21247 |
| Race and ethnicity | Mexican American | 1·00 (0·90–1·11) | 0·987 | 10460 | 6484 |
| Race and ethnicity | Other Hispanic | 0·96 (0·79–1·17) | 0·706 | 5725 | 3538 |
| Race and ethnicity | Non-Hispanic White | 0·89 (0·83–0·95) | <0·001 | 30141 | 20289 |
| Race and ethnicity | Non-Hispanic Black | 1·01 (0·91–1·13) | 0·799 | 13672 | 9544 |
| Race and ethnicity | Non-Hispanic Asian | 0·99 (0·69–1·41) | 0·938 | 3693 | 2033 |
| Race and ethnicity | Other/multiracial | 0·96 (0·78–1·20) | 0·742 | 2862 | 1808 |

Models were adjusted for age, sex, and race and ethnicity as applicable.

#### Supplementary Table 6A: Model comparison for exploratory one-knot trend screening

| Outcome | Linear AIC | Best knot year | One-knot AIC | ΔAIC | Preferred model |
|---|---|---|---|---|---|
| Adiposity criterion | -30·60 | 2016 | -33·80 | -3·20 | One-knot |
| CKM stages 2-4 | -31·01 | 2010 | -31·96 | -0·95 | Linear/inconclusive |
| Kidney risk | -21·66 | 2004 | -21·86 | -0·20 | Linear/inconclusive |
| Clinical cardiovascular disease | -25·81 | 2014 | -27·75 | -1·95 | Linear/inconclusive |
| Diabetes | -32·82 | 2016 | -34·29 | -1·48 | Linear/inconclusive |
| Hypertriglyceridemia | 4·84 | 2016 | 3·43 | -1·41 | Linear/inconclusive |
| Hypertension | -35·17 | 2008 | -35·17 | 0·00 | Linear/inconclusive |
| CKM stages 3-4 | -31·83 | 2012 | -32·55 | -0·72 | Linear/inconclusive |
| CKM stage 4 | -25·81 | 2014 | -27·75 | -1·95 | Linear/inconclusive |

#### Supplementary Table 6B: Estimated logit slopes from linear and one-knot models

| Outcome | Linear slope/year | Pre-knot slope/year | Post-knot slope/year |
|---|---|---|---|
| Adiposity criterion | 0·0275 | 0·0315 | 0·0044 |
| CKM stages 2-4 | -0·0060 | -0·0134 | -0·0004 |
| Kidney risk | -0·0007 | 0·0283 | -0·0050 |
| Clinical cardiovascular disease | -0·0046 | -0·0114 | 0·0092 |
| Diabetes | 0·0230 | 0·0268 | 0·0088 |
| Hypertriglyceridemia | -0·0231 | -0·0384 | 0·0296 |
| Hypertension | -0·0048 | -0·0122 | -0·0018 |
| CKM stages 3-4 | -0·0056 | -0·0114 | 0·0012 |
| CKM stage 4 | -0·0046 | -0·0114 | 0·0092 |

A one-knot model was considered preferred when AIC was more than 2 points lower than the linear model. Slopes are changes in log odds per calendar year. These analyses were hypothesis-generating.

#### Supplementary Table 7: Projection inputs and scenario equations

| Input or scenario | Specification | Interpretation |
|---|---|---|
| Adult population, 2023 | 254,244,076 | US Census 2023 National Population Projections |
| Adult population, 2040 | 279,789,047 | US Census 2023 National Population Projections |
| Adult population, 2050 | 286,248,739 | US Census 2023 National Population Projections |
| Age groups | 20–44, 45–64, ≥65 years | Matched between recent prevalence and projection composition |
| Population ageing only | Σa N(a,y) × p(a,2021–2023) | Age-specific prevalence held constant |
| Trend continuation | logit[p(a,y)] = logit[p(a,2021–2023)] + β × (y–2023)/10 | β from adjusted historical stage 2–4 model |
| Risk improvement | 0·90 × ageing-only prevalence in 2040; 0·80 × ageing-only prevalence in 2050 | Prespecified counterfactual scenario |
| Uncertainty | 20,000 Monte Carlo draws | Age-specific covariance and trend-coefficient uncertainty propagated |

a denotes age group; N denotes projected population; p denotes prevalence; β denotes the adjusted log-odds trend per decade.

#### Supplementary Table 8: Historical calibration of the stage 2–4 projection model

| Cycle | Midyear | Observed, % | Fitted, % | Fitted minus observed, pp |
|---|---|---|---|---|
| 1999-2000 | 2000.0 | 65·75 | 64·54 | -1·20 |
| 2001-2002 | 2002.0 | 63·11 | 64·17 | 1·06 |
| 2003-2004 | 2004.0 | 64·24 | 63·79 | -0·45 |
| 2005-2006 | 2006.0 | 63·48 | 63·41 | -0·06 |
| 2007-2008 | 2008.0 | 63·40 | 63·03 | -0·37 |
| 2009-2010 | 2010.0 | 60·56 | 62·65 | 2·09 |
| 2011-2012 | 2012.0 | 63·17 | 62·27 | -0·90 |
| 2013-2014 | 2014.0 | 62·56 | 61·88 | -0·68 |
| 2015-2016 | 2016.0 | 62·23 | 61·49 | -0·73 |
| 2017-March2020 | 2018.6 | 60·35 | 60·99 | 0·64 |
| 2021-2023 | 2022.0 | 62·13 | 60·32 | -1·80 |

The maximum absolute fitted–observed difference was 2·09 percentage points.

#### Supplementary Table 9: Sensitivity analyses

| Analysis | Outcome | Cycle | N | Prevalence, % |
|---|---|---|---|---|
| Primary maximum-observed staging | Any CKM | 1999-2000 | 4880 | 84·3 |
| Primary maximum-observed staging | CKM stages 2-4 | 1999-2000 | 4880 | 65·7 |
| Primary maximum-observed staging | CKM stages 3-4 | 1999-2000 | 4880 | 12·5 |
| Primary maximum-observed staging | Any CKM | 2021-2023 | 7809 | 88·5 |
| Primary maximum-observed staging | CKM stages 2-4 | 2021-2023 | 7809 | 62·1 |
| Primary maximum-observed staging | CKM stages 3-4 | 2021-2023 | 7809 | 11·7 |
| Laboratory-weighted staging | Any CKM | 1999-2000 | 4880 | 84·3 |
| Laboratory-weighted staging | CKM stages 2-4 | 1999-2000 | 4880 | 65·7 |
| Laboratory-weighted staging | CKM stages 3-4 | 1999-2000 | 4880 | 12·5 |
| Laboratory-weighted staging | Any CKM | 2021-2023 | 6064 | 89·1 |
| Laboratory-weighted staging | CKM stages 2-4 | 2021-2023 | 6064 | 62·5 |
| Laboratory-weighted staging | CKM stages 3-4 | 2021-2023 | 6064 | 11·9 |
| Exclude PREVENT predictor clipping | Any CKM | 1999-2000 | 4880 | 84·3 |
| Exclude PREVENT predictor clipping | CKM stages 2-4 | 1999-2000 | 4880 | 65·7 |
| Exclude PREVENT predictor clipping | CKM stages 3-4 | 1999-2000 | 4880 | 12·5 |
| Exclude PREVENT predictor clipping | Any CKM | 2021-2023 | 7176 | 87·9 |
| Exclude PREVENT predictor clipping | CKM stages 2-4 | 2021-2023 | 7176 | 60·4 |
| Exclude PREVENT predictor clipping | CKM stages 3-4 | 2021-2023 | 7176 | 12·3 |

Sensitivity estimates are age-standardised point estimates; primary design-based CIs are reported in the main analysis.

#### Supplementary Table 10: Complete-case and alternative-definition endpoint estimates

| Analysis | Outcome | Cycle | N | Age-standardised prevalence, % (95% CI) |
|---|---|---|---|---|
| Primary maximum-observed | Any CKM | 1999-2000 | 4,880 | 84·3 (82·2–86·3) |
| Primary maximum-observed | CKM stages 2–4 | 1999-2000 | 4,880 | 65·7 (63·0–68·5) |
| Primary maximum-observed | CKM stages 3–4 | 1999-2000 | 4,880 | 12·5 (10·9–14·2) |
| Primary maximum-observed | Any CKM | 2021-2023 | 7,809 | 88·5 (87·2–89·9) |
| Primary maximum-observed | CKM stages 2–4 | 2021-2023 | 7,809 | 62·1 (59·9–64·4) |
| Primary maximum-observed | CKM stages 3–4 | 2021-2023 | 7,809 | 11·7 (10·8–12·7) |
| Complete-case core variables | Any CKM | 1999-2000 | 1,904 | 86·1 (83·3–88·9) |
| Complete-case core variables | CKM stages 2–4 | 1999-2000 | 1,904 | 72·3 (69·3–75·3) |
| Complete-case core variables | CKM stages 3–4 | 1999-2000 | 1,904 | 10·6 (9·1–12·1) |
| Complete-case core variables | Any CKM | 2021-2023 | 2,815 | 89·5 (87·7–91·2) |
| Complete-case core variables | CKM stages 2–4 | 2021-2023 | 2,815 | 65·4 (62·7–68·2) |
| Complete-case core variables | CKM stages 3–4 | 2021-2023 | 2,815 | 11·5 (10·3–12·7) |
| PREVENT threshold 15% | CKM stages 3–4 | 1999-2000 | 4,880 | 15·7 (14·0–17·3) |
| PREVENT threshold 15% | CKM stages 3–4 | 2021-2023 | 7,809 | 14·4 (13·4–15·3) |
| PREVENT threshold 25% | CKM stages 3–4 | 1999-2000 | 4,880 | 10·7 (9·2–12·2) |
| PREVENT threshold 25% | CKM stages 3–4 | 2021-2023 | 7,809 | 10·2 (9·2–11·1) |
| Uniform adiposity thresholds | Any CKM | 1999-2000 | 4,880 | 84·3 (82·2–86·3) |
| Uniform adiposity thresholds | CKM stages 2–4 | 1999-2000 | 4,880 | 66·5 (63·6–69·5) |
| Uniform adiposity thresholds | CKM stages 3–4 | 1999-2000 | 4,880 | 12·5 (10·9–14·2) |
| Uniform adiposity thresholds | CKM stage 1 | 1999-2000 | 4,880 | 17·7 (15·4–20·1) |
| Uniform adiposity thresholds | Any CKM | 2021-2023 | 7,809 | 88·1 (86·6–89·6) |
| Uniform adiposity thresholds | CKM stages 2–4 | 2021-2023 | 7,809 | 62·6 (60·3–64·9) |
| Uniform adiposity thresholds | CKM stages 3–4 | 2021-2023 | 7,809 | 11·7 (10·8–12·7) |
| Uniform adiposity thresholds | CKM stage 1 | 2021-2023 | 7,809 | 25·5 (23·9–27·0) |

Complete-case analyses used laboratory-specific weights. Alternative PREVENT thresholds affect stage 3 assignment; uniform adiposity thresholds removed Asian-specific lower body-mass index and waist cut points. Estimates are directly age standardised to the 2010 US adult population.

#### Supplementary Table 11: Sensitivity analyses of long-term trends

| Analysis | Outcome | Adjusted OR per decade (95% CI) | P | N | Events |
|---|---|---|---|---|---|
| Complete-case core variables | Any CKM | 1·11 (1·01–1·23) | 0·026 | 25,680 | 23,332 |
| Complete-case core variables | CKM stages 2–4 | 0·81 (0·76–0·87) | <0·001 | 25,680 | 18,451 |
| Complete-case core variables | CKM stages 3–4 | 0·96 (0·88–1·04) | 0·305 | 25,680 | 4,192 |
| PREVENT threshold 15% | CKM stages 3–4 | 0·94 (0·89–1·00) | 0·041 | 66,553 | 13,450 |
| PREVENT threshold 15% | CKM stage 3 | 0·99 (0·93–1·06) | 0·875 | 66,553 | 5,645 |
| PREVENT threshold 25% | CKM stages 3–4 | 0·96 (0·90–1·03) | 0·284 | 66,553 | 9,423 |
| PREVENT threshold 25% | CKM stage 3 | 1·01 (0·91–1·12) | 0·914 | 66,553 | 1,618 |
| Uniform adiposity thresholds | Any CKM | 1·23 (1·15–1·31) | <0·001 | 66,553 | 57,028 |
| Uniform adiposity thresholds | CKM stages 2–4 | 0·93 (0·88–0·98) | 0·010 | 66,553 | 44,075 |
| Uniform adiposity thresholds | CKM stages 3–4 | 0·95 (0·89–1·01) | 0·102 | 66,553 | 10,998 |
| Uniform adiposity thresholds | CKM stage 1 | 1·22 (1·16–1·28) | <0·001 | 66,553 | 12,953 |
| Exclude 2021-2023 period | Any CKM | 1·26 (1·15–1·37) | <0·001 | 58,744 | 50,984 |
| Exclude 2021-2023 period | CKM stages 2–4 | 0·90 (0·84–0·97) | 0·005 | 58,744 | 38,768 |
| Exclude 2021-2023 period | CKM stages 3–4 | 0·96 (0·89–1·04) | 0·328 | 58,744 | 9,695 |
| Exclude 2021-2023 period | CKM stage 0 | 0·80 (0·73–0·87) | <0·001 | 58,744 | 7,760 |
| Exclude 2021-2023 period | CKM stage 1 | 1·28 (1·21–1·36) | <0·001 | 58,744 | 12,216 |

Odds ratios are per decade and account for NHANES weights, strata, and primary sampling units. Complete-case analyses used laboratory-specific weights. Models excluding August 2021–August 2023 ended with the 2017–March 2020 period.

**Supplementary Table 12: Socioeconomic gradients in CKM stages 2–4, August 2021–August 2023**

| Dimension | Level | N | Age-standardised prevalence, % (95% CI) | Adjusted OR (95% CI) | P |
|---|---|---|---|---|---|
| Education | College or above | 2,625 | 54·4 (50·8–57·9) | 1·00 (reference) | Reference |
| Education | Some college | 2,370 | 64·7 (62·4–67·1) | 1·68 (1·33–2·12) | <0·001 |
| Education | High school/GED | 1,749 | 66·6 (62·5–70·8) | 1·85 (1·38–2·48) | <0·001 |
| Education | <High school | 1,039 | 69·6 (64·6–74·6) | 2·17 (1·60–2·94) | <0·001 |
| Family income-to-poverty ratio | High income | 2,572 | 58·5 (54·9–62·2) | 1·00 (reference) | Reference |
| Family income-to-poverty ratio | Middle income | 2,446 | 64·3 (61·4–67·2) | 1·38 (1·11–1·71) | 0·006 |
| Family income-to-poverty ratio | Low income | 1,471 | 67·0 (64·1–69·9) | 1·56 (1·31–1·87) | <0·001 |
| Insurance | Insured | 7,108 | 62·0 (59·6–64·3) | 1·00 (reference) | Reference |
| Insurance | Uninsured | 668 | 64·1 (59·1–69·2) | 1·02 (0·81–1·30) | 0·834 |

Prevalence estimates are directly age standardised. Odds ratios are from separate survey-weighted logistic models adjusted for age, sex, and race and ethnicity. Reference groups were college or above, high income, and insured. These analyses are exploratory.

#### Supplementary Table 13: Rolling-origin temporal validation of the adjusted stages 2–4 trend model

| Model trained through | Validation cycle | Observed, % | Predicted, % | Prediction error, pp |
|---|---|---|---|---|
| 2007-2008 | 2009-2010 | 60·56 | 62·75 | +2·19 |
| 2009-2010 | 2011-2012 | 63·17 | 60·52 | -2·64 |
| 2011-2012 | 2013-2014 | 62·56 | 61·29 | -1·27 |
| 2013-2014 | 2015-2016 | 62·23 | 61·81 | -0·42 |
| 2015-2016 | 2017-March2020 | 60·35 | 61·74 | +1·38 |
| 2017-March2020 | 2021-2023 | 62·13 | 60·52 | -1·60 |

Across six sequential validation cycles, mean absolute error was 1·58 percentage points, root mean squared error was 1·73 points, and maximum absolute error was 2·64 points. In the post-2016 holdout analysis, absolute errors were 1·38 points for 2017–March 2020 and 0·89 points for 2021–2023.

#### Supplementary Table 14: Unweighted comparison of the full analytic cohort and complete-case subset

| Characteristic | Full analytic cohort | Complete-case subset | Standardised difference |
|---|---|---|---|
| Participants, n | 66,553 | 25,680 | — |
| Age, years | 50·4 (18·3) | 50·0 (17·8) | −0·023 |
| Body-mass index, kg/m² | 29·1 (6·9) | 28·9 (6·6) | −0·024 |
| Women | 52·3% | 51·6% | −0·015 |
| Mexican American | 15·7% | 16·4% | 0·019 |
| Other Hispanic | 8·6% | 8·9% | 0·011 |
| Non-Hispanic White | 45·3% | 46·3% | 0·020 |
| Non-Hispanic Black | 20·5% | 18·9% | −0·042 |
| Non-Hispanic Asian | 5·5% | 5·5% | −0·002 |
| Other/multiracial | 4·3% | 4·0% | −0·015 |
| Diabetes | 17·8% | 17·5% | −0·006 |
| Clinical cardiovascular disease | 10·6% | 9·6% | −0·031 |
| CKM stages 2–4 | 65·7% | 71·8% | 0·134 |
| Survey period: 2017–March 2020 | 13·9% | 13·3% | −0·018 |
| Survey period: 2021–2023 | 11·7% | 11·0% | −0·024 |

Values are unweighted mean (SD) or proportion. Standardised differences compare the complete-case subset with the full analytic cohort. Absolute values below 0·10 indicate little imbalance; the complete-case subset was modestly enriched for CKM stages 2–4. Survey-period standardised differences for all earlier period groups were below 0·03.

#### Supplementary Figures

**Supplementary Figure 1: Age-standardised prevalence of any CKM, stages 2–4, and stages 3–4 across NHANES periods.** Error bars indicate 95% CIs.

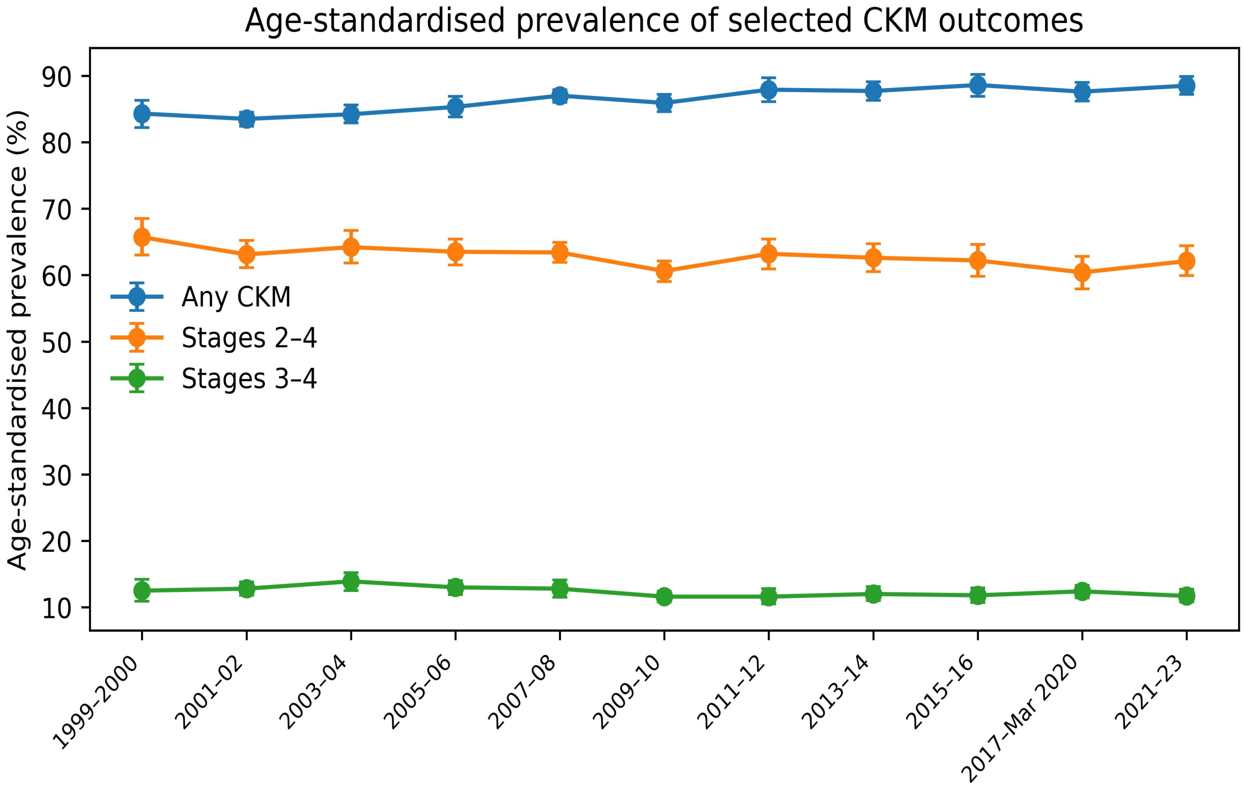

**Supplementary Figure 2: Observed versus model-fitted stage 2–4 prevalence used for historical calibration.**

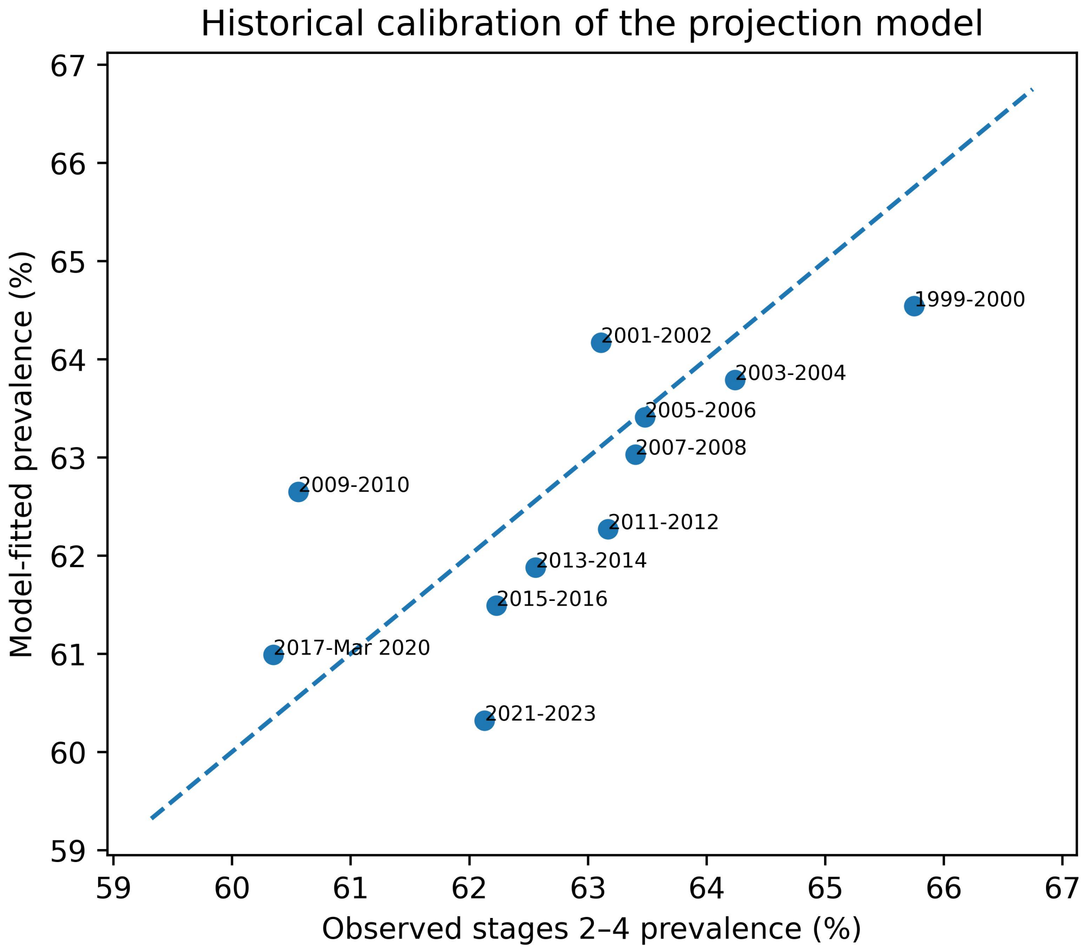

**Supplementary Figure 3: Endpoint prevalence estimates in 2021–2023 under the primary staging, laboratory-weighted analysis, and exclusion of PREVENT predictor truncation.**

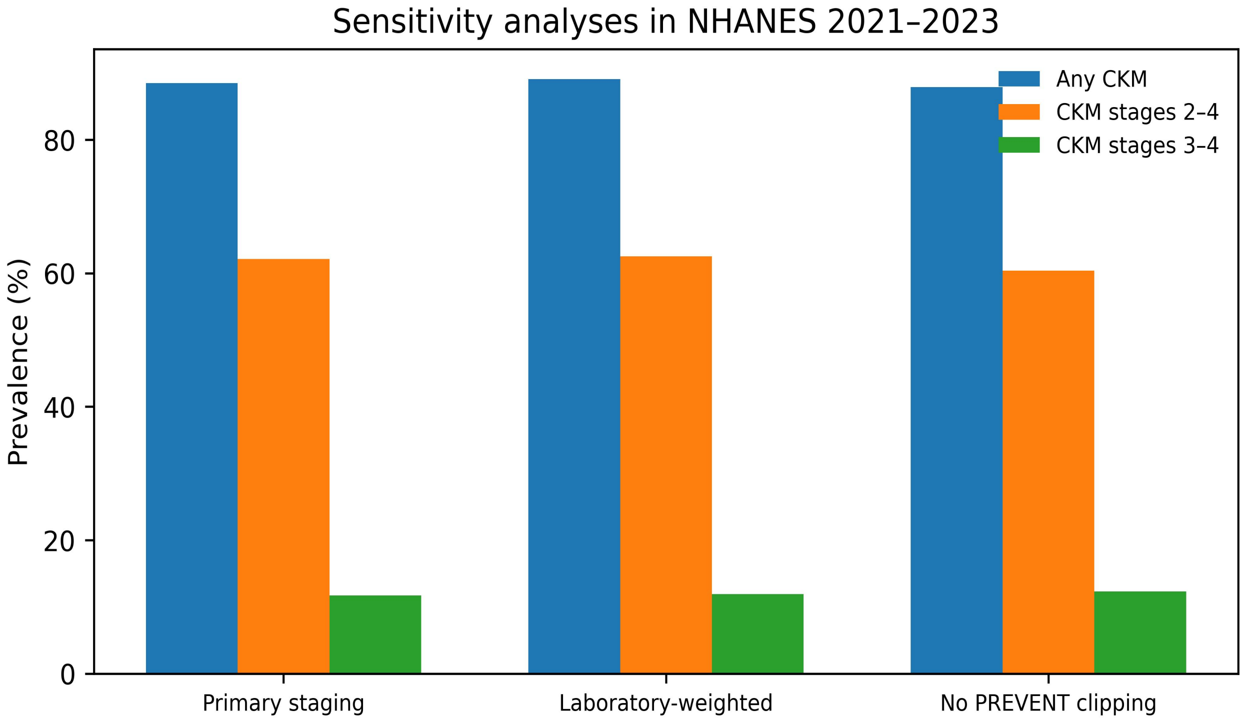

**Supplementary Figure 4: Adjusted trends in CKM stages 2–4 by sex, age, and race and ethnicity.** Odds ratios are per decade from survey-weighted logistic models adjusted for age, sex, and race and ethnicity as applicable. Error bars indicate 95% CIs.

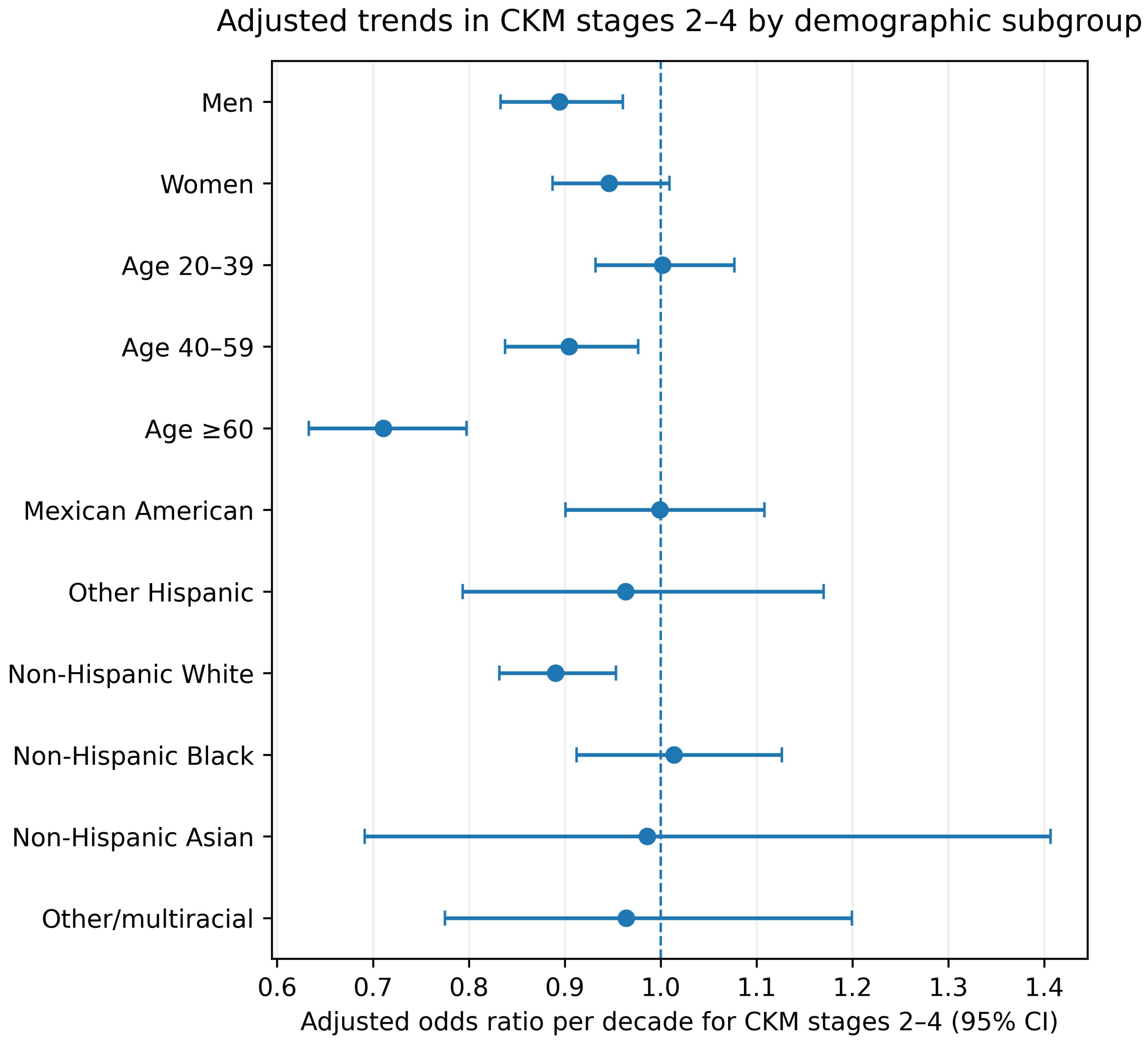
